# Effectiveness of COVID-19 Vaccines among Incarcerated People in California State Prisons: A Retrospective Cohort Study

**DOI:** 10.1101/2021.08.16.21262149

**Authors:** Elizabeth T. Chin, David Leidner, Yifan Zhang, Elizabeth Long, Lea Prince, Stephanie J. Schrag, Jennifer R. Verani, Ryan E. Wiegand, Fernando Alarid-Escudero, Jeremy D. Goldhaber-Fiebert, David M. Studdert, Jason R. Andrews, Joshua A. Salomon

## Abstract

**Background:** Prisons and jails are high-risk settings for COVID-19 transmission, morbidity, and mortality. COVID-19 vaccines may substantially reduce these risks, but evidence is needed of their effectiveness for incarcerated people, who are confined in large, risky congregate settings.

**Methods:** We conducted a retrospective cohort study to estimate effectiveness of mRNA vaccines, BNT162b2 (Pfizer-BioNTech) and mRNA-1273 (Moderna), against confirmed SARS-CoV-2 infections among incarcerated people in California prisons from December 22, 2020 through March 1, 2021. The California Department of Corrections and Rehabilitation provided daily data for all prison residents including demographic, clinical, and carceral characteristics, as well as COVID-19 testing, vaccination status, and outcomes. We estimated vaccine effectiveness using multivariable Cox models with time-varying covariates that adjusted for resident characteristics and infection rates across prisons.

**Findings:** Among 60,707 residents in the cohort, 49% received at least one BNT162b2 or mRNA-1273 dose during the study period. Estimated vaccine effectiveness was 74% (95% confidence interval [CI], 64−82%) from day 14 after first dose until receipt of second dose and 97% (95% CI, 88−99%) from day 14 after second dose. Effectiveness was similar among the subset of residents who were medically vulnerable (74% [95% CI, 62−82%] and 92% [95% CI, 74−98%] from 14 days after first and second doses, respectively), as well as among the subset of residents who received the mRNA-1273 vaccine (71% [95% CI, 58−80%] and 96% [95% CI, 67−99%]).

**Conclusions:** Consistent with results from randomized trials and observational studies in other populations, mRNA vaccines were highly effective in preventing SARS-CoV-2 infections among incarcerated people. Prioritizing incarcerated people for vaccination, redoubling efforts to boost vaccination and continuing other ongoing mitigation practices are essential in preventing COVID-19 in this disproportionately affected population.

**Funding:** Horowitz Family Foundation, National Institute on Drug Abuse, Centers for Disease Control and Prevention, National Science Foundation, Open Society Foundation, Advanced Micro Devices.

## Introduction

The BNT162b2 (Pfizer-BioNTech) and mRNA-1273 (Moderna) vaccines appear highly effective in preventing severe acute respiratory syndrome coronavirus 2 (SARS-CoV-2) infection and coronavirus disease 2019 (COVID-19) illness. Augmenting efficacy evidence from clinical trials,^1,2^ observational studies among healthcare workers,^3,4^ adults aged 65 years or older,^5^ and the general community^6,7^ have reported levels of protection from full vaccination ranging from 89% to 95%. However, except for two relatively small studies of partial vaccination in skilled nursing facilities^8,9^, no published studies to date have examined the effectiveness of COVID-19 vaccines in congregate settings, where risks of transmission are very high.

Prisons and jails are especially risky congregate settings. Living quarters are often densely populated and poorly ventilated, physical distancing is typically infeasible, and pre-existing medical conditions associated with severe COVID-19 illness are prevalent among incarcerated people.^10,11^ Recognizing these risks and the considerable potential for vaccines to reduce them, approximately half of states in the USA have prioritized incarcerated people for COVID-19 vaccines. In contrast, 15 states have not included incarcerated people in vaccine distribution plans or have assigned them to lowest priority tiers.^12,13^

The California Department of Corrections and Rehabilitation (CDCR), which operates the second largest state prison system in the USA, launched a COVID-19 vaccination program on December 22, 2020, and rapidly scaled up the program across its 35 prisons.^14^ CDCR has conducted extensive testing and collected detailed data relevant to COVID-19 risks, interventions, and outcomes. We analyzed these data to estimate effectiveness of mRNA vaccines against confirmed SARS-CoV-2 infection among nearly 61,000 incarcerated people in California.

## Methods

### Study design and population

We conducted a retrospective cohort study spanning the 70-day period from December 22, 2020, through March 1, 2021, during which residents were offered either BNT162b2 or mRNA-1273 vaccines. Prioritization criteria CDCR used to direct first-dose offers changed over time as supply expanded and state and federal guidance evolved. Criteria included residency in a specialized medical or psychiatric care setting, age and medical comorbidities, no confirmed SARS-CoV-2 infection (or none in the previous 90 days), and participation in penal labor. CDCR prioritized timely second-dose offers to adhere to recommended dosing schedules.

Residents were eligible for inclusion in the study cohort if they were incarcerated in a CDCR prison on the study start date and had no prior confirmed SARS-CoV-2 infection. Cohort members contributed observation time beginning on the study start date and ending on the day of the earliest of the following events: release from CDCR custody, sample collection for a positive SARS-CoV-2 diagnostic test, or study end date.

### Data and key measures

CDCR collects and stores daily data on each resident. Data provided for this study included demographic characteristics (sex, age, racial or ethnic group), documented history of 25 comorbid conditions (e.g., hypertension, chronic kidney disease, asthma), and a composite COVID-19 risk score. CDCR designed the COVID-19 risk score to grade risks of severe illness from SARS-CoV-2 infections based on individual demographic and clinical information (Table S1), and the agency has used this score to guide COVID-19 mitigation policies, including prioritization of testing and vaccination. We also obtained person-day level variables indicating each resident’s prison, facility, building, housing unit, floor, and room of residence; room type (cell or dormitory); security level; and participation in penal labor.

Detailed SARS-CoV-2 testing information came from a multilayered resident testing program that included risk-based routine testing, surveillance testing, and testing in response to detected outbreaks (Table S2). Information provided on accepted vaccine doses allowed us to classify cohort members’ daily vaccination status into six categories: unvaccinated, from 0 to 6 days after receiving a first dose, from 7 to 13 days after a first dose, from 14 days after a first dose until receipt of a second dose, from 0 to 13 days after a second dose, and from 14 days after a second dose.

To obtain a measure of risk of infection from correctional staff, we constructed a prison-day level variable comprising the rolling 7-day COVID-19 case rate among staff at each prison. Infections among correctional staff were identified through a program of regular SARS-CoV-2 testing, mandated and administered by CDCR (Table S2).

### Statistical analysis

To obtain estimates of vaccine effectiveness, we fit multivariable models using the Andersen–Gill extension of the Cox proportional hazards model^15^ to account for time-varying covariates using person-day level data. The primary outcome of interest was SARS-CoV-2 infection, confirmed by positive PCR or antigen test. We specified exposure status according to the six vaccination categories described above. Effectiveness estimates are expressed as 1 minus the hazard ratio.

Analyses adjusted for residents’ racial or ethnic group, COVID-19 risk score, security level, room type, participation in penal labor, staff case rate, and prison (fixed effects). We did not adjust for sex because men and women are generally housed in separate prisons, making this variable highly collinear with prison. To account for non-independence between cohort members, we clustered standard errors by housing unit. Housing units are discrete cohorts within prisons, consisting of residents who co-participate in activities (e.g., recreation, laundry, dining).

All analyses were performed using R software, version 3.5.2 (R Foundation for Statistical Computing). Additional details regarding model and variable specifications are provided in the Supplementary Appendix.

### Secondary analyses

We conducted four sets of secondary analyses. First, we estimated effectiveness in two subgroups of interest. Specifically, recognizing that our primary analysis mixes effects of two different vaccines, we ran one subgroup analysis focusing on mRNA-1273 vaccinations only (which accounted for 78% of all first doses and 72% of all doses administered in the study period). We also estimated effectiveness among medically vulnerable residents by restricting the analytic cohort to residents with COVID-19 risk scores of 2 or higher, indicating moderate or high risk. Residents with COVID-19 risk scores of 2 or higher were either aged 65 years and older or younger than 65 years with comorbid conditions associated with severe COVID-19 disease (Table S1).

Second, we estimated effectiveness in a broader population that included residents with prior infections and those who entered prison during the study period. Third, we examined the sensitivity of our effectiveness estimates to alternative model specifications, including censoring observation time at the collection date of cohort members’ last test (to exclude time periods in which infection status was unknown), and computing cluster-robust variance estimators with clusters defined at various levels (prison, facility, building, housing unit, floor, room, and person). Finally, to assess the sensitivity of estimates to choice of study period, we re-estimated effectiveness using a series of alternative study end dates between February 15 and July 1, 2021.

### Study oversight

The study was approved by the institutional review board at Stanford University (protocol #55835). It was reviewed by CDC and conducted according to applicable federal law and CDC policy.^*^ Results are reported in accordance with Strengthening the Reporting of Observational Studies in Epidemiology (STROBE) guidelines (checklist in Supplementary Appendix).^16^

## Results

### Sample characteristics and vaccination uptake

60,707 residents met the cohort inclusion criteria (Figure S1) and were followed for an average of 57.6 days (median, 70 days). By February 1, 2021, 20% of them received at least one mRNA dose and 3% received two doses; by March 1, 49% received at least one dose and 22% received two doses (Figure 1). The mean interval between doses was 20.8 days (standard deviation [SD]: 2.7) for those who received two BNT162b2 doses and 28.0 days (SD: 3.5) days for those who received two mRNA-1273 doses.

**Figure 1.**
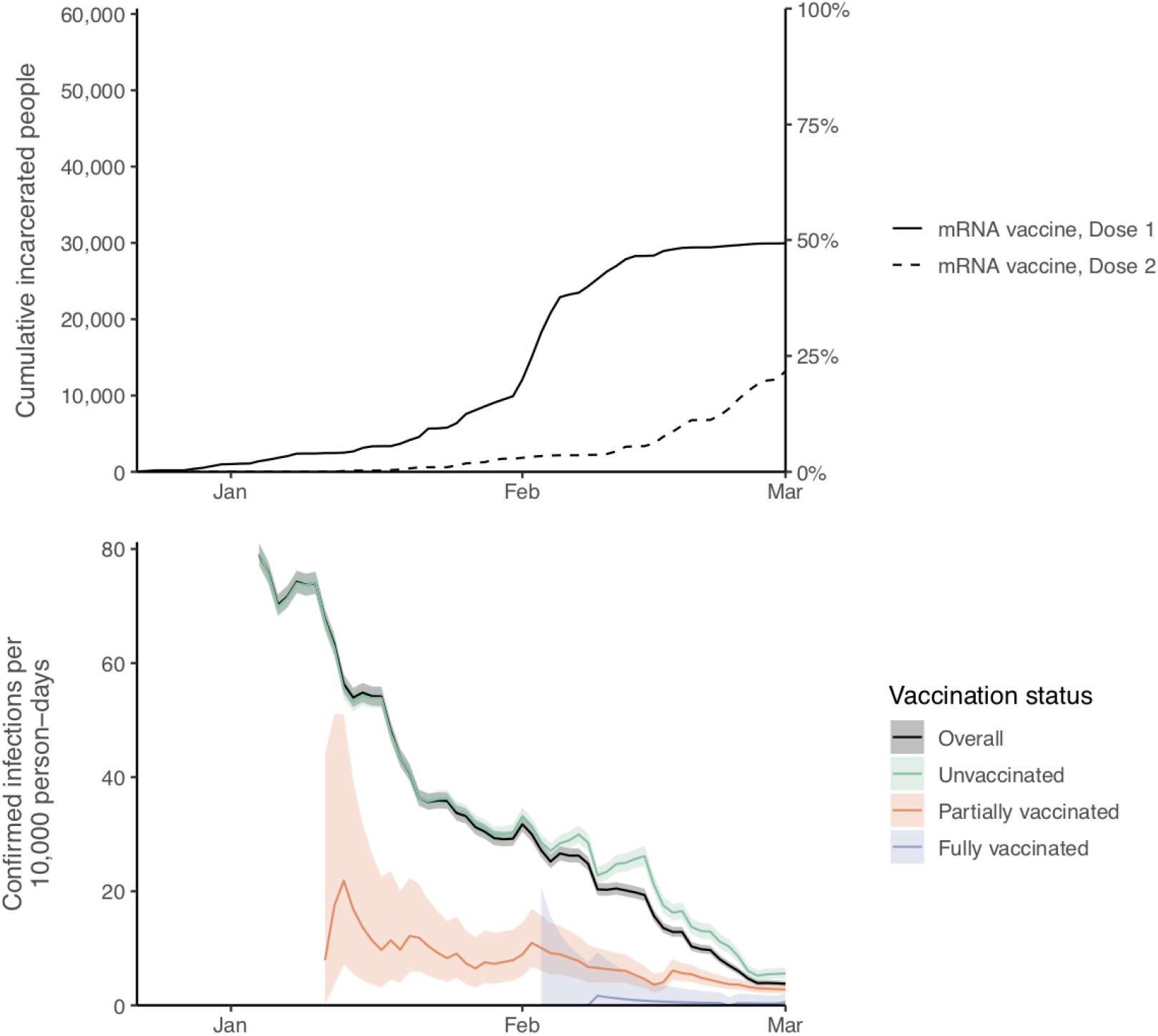
Cumulative vaccinations with one or two doses of mRNA vaccines (top panel) and 14-day rolling rates of confirmed infections per 10,000 person-days by vaccination status (bottom panel), among study cohort of incarcerated people in California state prisons.^*^ Shaded areas represent 95% confidence intervals. Partially vaccinated status defined as ≥14 days after a first dose until receipt of a second dose; fully vaccinated status defined as ≥14 days after a second dose. ^*^ Time periods with fewer than 200 people tested were excluded.

Most cohort members were male (96%), younger than 60 years (88%), and either Hispanic or Latino (43%) or non-Hispanic Black or African American (33%) (Table 1). Most had risk factors for severe outcomes from COVID-19 infection: 84% had at least one medical condition defined by CDC as a marker of severe COVID-19-related illness,^17^ and 31% had moderate or high COVID-19 risk according to CDCR’s scoring algorithm. Cohort members who had received one or more vaccine doses by the end of the study period tended to be older than those who had not, and were more likely to have medical conditions and higher COVID-19 risk scores and be non-Hispanic White or Hispanic or Latino (Table 1).

**Table 1.**
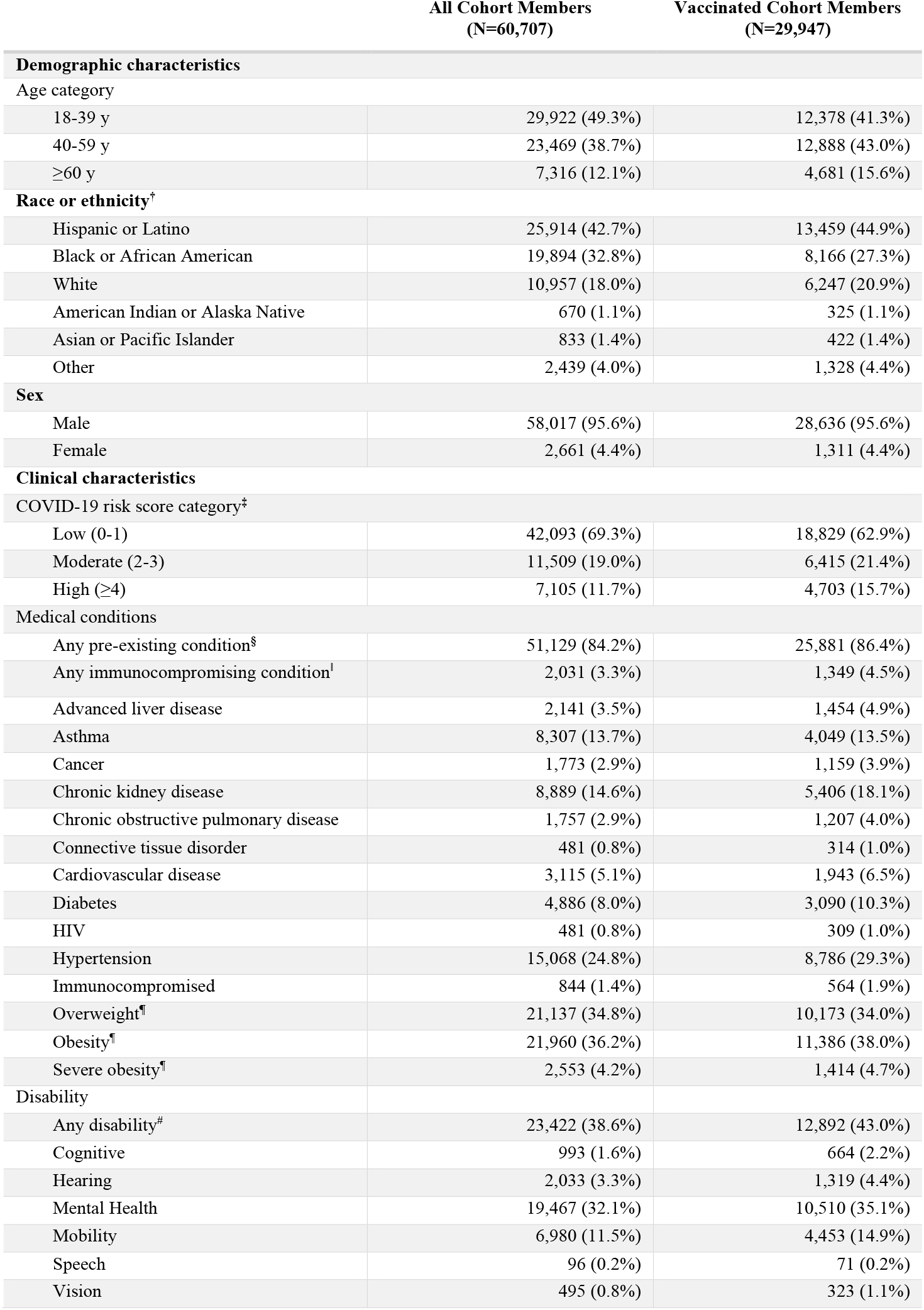
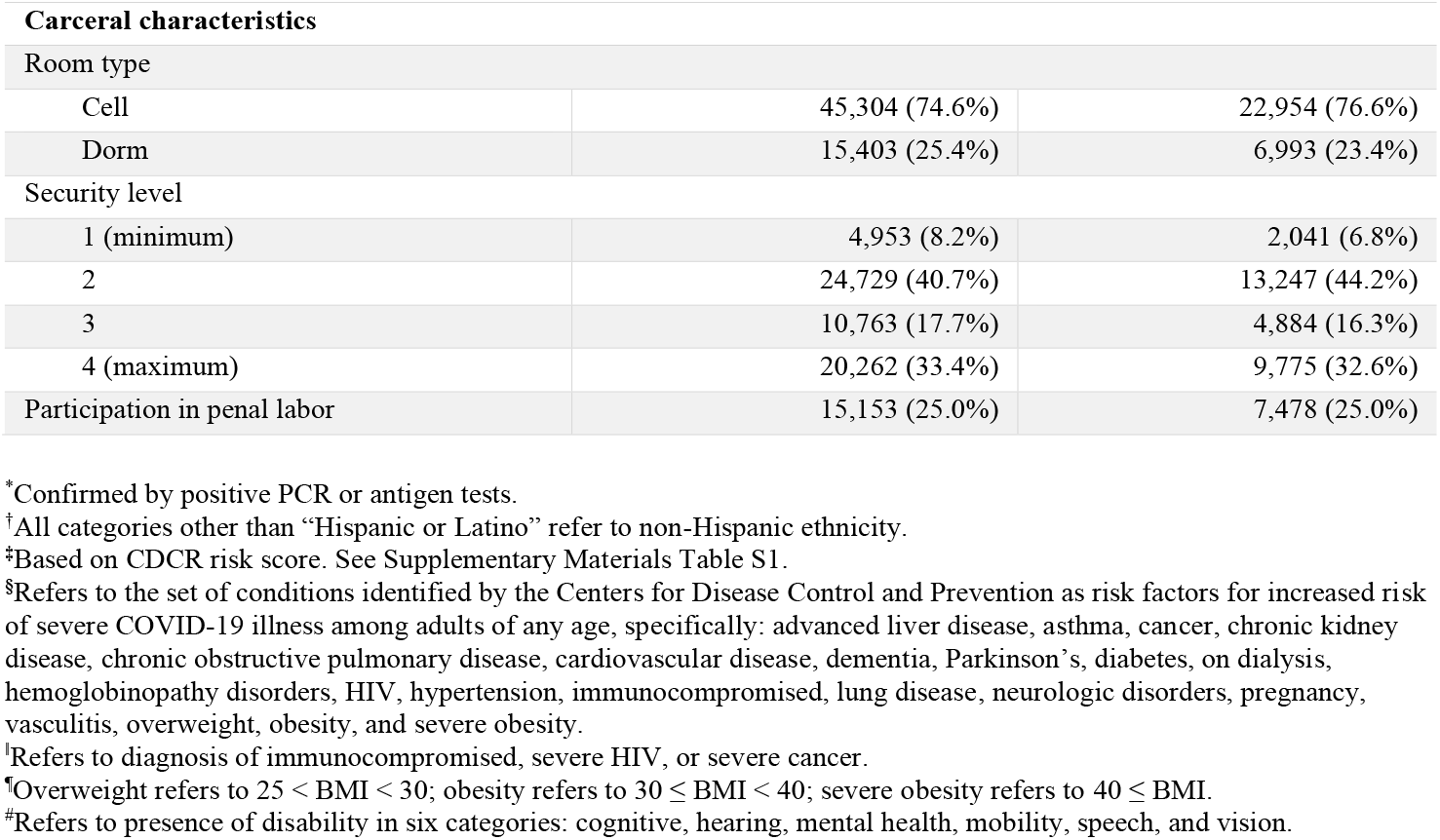
Demographic, health, and carceral characteristics of the study cohort of incarcerated people in California state prisons. Persons within the study cohort were incarcerated on December 22, 2020 and did not have prior confirmed SARS-CoV-2 infection^*^ documented in CDCR clinical records. Vaccinated residents were vaccinated between December 22, 2020 and March 1, 2021.

### Testing rates by vaccination category

Cohort members had a median of 6 COVID-19 tests during the study period (interquartile range: 2-10). Testing rates were lower in the unvaccinated group (Figure S2). In January 2021, for example, there were 933 tests per 10,000 person-days among the unvaccinated group, compared with 1,167 among the partially vaccinated group (≥14 days after first dose until receipt of second dose) and 2,018 among the fully vaccinated group (≥14 days after second dose). The rate of testing decreased from 957 tests per 10,000 person-days in January to 886 in February.

### Confirmed infections and other COVID-19 outcomes

A total of 13,216 confirmed infections (37.8 per 10,000 person-days), 393 hospitalizations (1.1 per 10,000 person-days), and 48 deaths (0.1 per 10,000 person-days) were documented among cohort members. Most of these outcomes occurred among unvaccinated people (Table 2). Incidence of confirmed infection was 0.6 per 10,000 person-days among the fully vaccinated, 3.5 among the partially vaccinated, and 46.8 among the unvaccinated. Incidence of infection decreased during the study period, from 40.2 per 10,000 person-days in January 2021 to 11.8 in February (Figure 1). Additional details on testing and confirmed infections, including time series by specific prison, are shown in Figures S2, S3, and S4 in the Supplementary Appendix.

**Table 2.** Persons, person-days, and vaccine effectiveness against COVID-19 infection among study cohort of incarcerated people in California state prisons, by vaccination status, December 22, 2020 to March 1, 2021.

| COVID-19 vaccination status | Confirmed infection* | Hospitalized<sup>†</sup> | Died<sup>‡</sup> | Tested | Total | Median follow-up, days | Person-days | Positive per 10,000 person-days | Effectiveness, Unadjusted<sup>§</sup> | Effectiveness, Adjusted<sup>§</sup> |
| --- | --- | --- | --- | --- | --- | --- | --- | --- | --- | --- |
| **Unvaccinated** | 12,318 | 356 | 44 | 53,415 | 60,673 | 43 | 2,633,734 | 46.8 | (Ref) | (Ref) |
| **Vaccinated with one dose** |  |  |  |  |  |  |  |  |  |  |
| 0-6 days after first dose | 527 | 20 | 3 | 19,767 | 29,947 | 7 | 206,960 | 25.5 | -4% (-44 to 25) | 16% (-15 to 39) |
| 7-13 days after first dose | 237 | 11 | 1 | 17,200 | 28,902 | 7 | 199,746 | 11.9 | 26% (-8 to 50) | 44% (20 to 61) |
| ≥14 days after first dose until second dose | 101 | 4 | 0 | 16,436 | 27,392 | 11 | 286,856 | 3.5 | 63% (48 to 74) | 74% (64 to 82) |
| **Vaccinated with two doses** |  |  |  |  |  |  |  |  |  |  |
| 0-13 days after second dose | 30 | 2 | 0 | 7,152 | 13,183 | 11 | 120,141 | 2.5 | 74% (41 to 89) | 85% (66 to 94) |
| ≥14 days after second dose | 3 | 0 | 0 | 2381 | 3,659 | 14 | 50,033 | 0.6 | 93% (76 to 98) | 97% (88 to 99) |

\*Confirmed SARS-CoV-2 infection is defined as having a positive PCR or antigen diagnostic test for SARS-CoV-2.
<sup>†</sup>Hospitalization related to a SARS-CoV-2 infection is defined as a hospitalization that occurred within three days prior to or 14 days after an infection was initially confirmed. For attribution to person-days stratified by vaccination category, hospitalizations were assigned to the collection date for a confirmed infection.
<sup>‡</sup>All deaths related to a SARS-CoV-2 infection were classified and confirmed by the California Correctional Health Care Services. For attribution to person-days stratified by vaccination category, deaths were assigned to the collection date for a confirmed infection.
<sup>§</sup>Unadjusted effectiveness estimates based on Cox proportional hazards model with only vaccination status indicators as explanatory variables. Adjusted effectiveness estimates based on Cox proportional hazards model including controls for residents' race or ethnic group (Hispanic, non-Hispanic Black or African American, non-Hispanic White, non-Hispanic American Indian or Alaska Native, non-Hispanic Asian or Pacific Islander, non-Hispanic Other), COVID-19 risk score (0 to ≥4, top-coded), security level (1, 2, 3, 4), room type (cell, dorm), involvement in penal labor (yes, no), the prison-specific 7-day rolling COVID-19 case rate for staff (continuous), and prison (fixed-effect).

### Vaccine effectiveness

There was no significant difference in the adjusted hazard ratio for confirmed SARS-CoV-2 infection during days 0 to 6 days after receiving a first dose relative to unvaccinated status (Table 2). From 7 to 13 days after a first dose, estimated vaccine effectiveness was 44% (95% CI, 20-61%), and from 14 days after a first dose until receipt of a second dose, effectiveness was 74% (95% CI, 64-82%). Effectiveness estimates were 85% (95% CI, 66-94%) from 0 to 13 days after a second dose and 97% (95% CI, 88-99%) from 14 days after a second dose.

### Secondary analyses

Subgroup analyses produced similar estimates of effectiveness to the full cohort analysis (Table S3A). Among those receiving the mRNA-1273 vaccine, estimated effectiveness was 71% (95% CI, 58-80%) from 14 days after first dose until receipt of second dose and 96% (95% CI, 67-99%) from 14 days after second dose. Among cohort members at moderate or high risk for severe COVID-19 illness, effectiveness estimates were 74% (95% CI, 62-82%) from 14 days after first dose until receipt of second dose and 92% (95% CI, 74-98%) from 14 days after second dose.

Estimates in an expanded cohort that included new entrants and residents with prior infections did not differ appreciably from the main cohort analysis (Table S3B). Results were also insensitive to model specification choices, including censoring of observation time at the date of cohort members’ last test and clustering standard errors at different residential levels (Table S3C and S3D).

In secondary analyses that modified the study end date, effectiveness estimates for fully vaccinated residents (i.e., from 14 days after second dose) decreased from 98% (95% CI, 82-100%) to 82% (95% CI, 69-89%) over a series of end dates between February 15 and July 1, 2021 (Table S3E). Study months spanning March to July were characterized by significantly lower outbreak risks across all facilities (0.4 confirmed infections per 10,000 person-days); lower testing (474 tests per 10,000 person-days); and high overall vaccination coverage rates (72% and 75% of cohort members who were still in custody had received at least one dose or had tested positive by April 1 and July 1, respectively).

## Discussion

This study found that BNT162b2 and mRNA-1273 vaccines were highly effective against confirmed SARS-CoV-2 infection among members of a high-risk and racially diverse population of incarcerated people. Beginning 14 days after a second mRNA vaccine dose, estimated effectiveness in this population was 97%. The vaccines were also highly effective among prison residents at higher risk for severe COVID-19 illness.

Our estimates of effectiveness among fully-vaccinated people in California prisons was higher than estimates reported by Cavanaugh et al^8^ from a skilled nursing facility in the USA (66% among residents and 76% among staff from 14 days after a second BNT162b2 dose), though similar to those reported for healthcare and other frontline workers by Thompson et al^3^ in the USA (91% from 14 days after second mRNA vaccine dose) and Angel et al^4^ in Israel (86% from 7 days after second BNT162b2 dose). Population-level studies in Israel by Dagan et al^7^ and Haas et al^6^ also reported similar results (92% and 95%, respectively, from 7 days after second BNT162b2 dose) as our study. Estimates of effectiveness of partial vaccination are more variable. We estimated 74% effectiveness against infection from 14 days after a first mRNA vaccine dose until receipt of second dose. This result was lower than Thompson et al’s^3^ estimate of 81% among healthcare and other frontline workers from 14 days after first mRNA vaccine dose until 14 days after second dose, but substantially higher than Dagan et al’s^7^ estimates of 46% for days 14 through 20 after first BNT162b2 dose and 60% for days 21 through 27.

To our knowledge, this is the first study to assess effectiveness of a COVID-19 vaccination program in a carceral setting. It has several strengths. We used detailed daily information on vaccination status and key COVID-19 outcomes for each resident. These data allowed us to adjust for key potential confounders, including risk factors for severe COVID-19, housing arrangements, and participation in penal labor. An extensive testing program in this population facilitated relatively complete measurement of SARS-CoV-2 infections. In addition, the large sample size permitted estimates of effectiveness within particular subgroups of interest (e.g., medically vulnerable).

Understanding vaccine effectiveness among people at high risk for severe disease is a priority. Our estimated effectiveness for partial and full vaccination did not differ appreciably between the full cohort and subsets characterized by moderate or high risk for severe COVID-19 illness. This bolsters growing evidence that mRNA vaccines provide substantial protection in older adults,^5,7^ people with pre-existing conditions,^7,18^ and residents of skilled nursing facilities.^8,9^ Our results also extend evidence from studies of healthcare workers indicating these vaccines are effective in environments characterized by high transmission risks.

In observational cohort studies like ours, potential for bias due to confounding is an important consideration. Vaccines were not offered randomly to residents—in particular those with risk factors for severe disease were prioritized. Given the latency of biologically plausible protection, the days after vaccination can serve as an indicator of bias, with large effectiveness estimates signaling substantial residual confounding.^19^ We included an exposure category for the first week after a first mRNA vaccine dose to assess the presence of such residual confounding, and detected a statistically insignificant 16% effectiveness for this negative control exposure. Vasileiou et al.^18^ reported a much higher estimate, 86% protection against COVID-19 hospitalizations during the first week after vaccination for BNT162b2, in a previous study on effectiveness in Scotland.

Residents were tested frequently (median 6 tests) during the 70-day study period, but testing was neither routine, random nor compulsory, creating potential for ascertainment bias. Several results provide some reassurance in this regard. First, vaccinated cohort members overall had 25% higher testing rates than unvaccinated members. Thus, the most plausible bias from differential testing would be more complete case detection among the vaccinated, which would lead to underestimating vaccine effectiveness. Second, an analysis that censored follow-up on the last test collection date for a cohort member produced effectiveness estimates similar to those from the main analysis.

Extending the study period through July 1, 2021 added four months in which testing and case rates were low, and a relatively large proportion of prison residents had been vaccinated. We found lower levels of estimated effectiveness for the fully vaccinated group over this extended period—an expected result, and a trend seen in the six-month vaccine efficacy clinical trial for the BNT162b2 vaccine.^20^ Accumulation of undetected infections that confer natural immunity may have contributed to dilution of estimated effectiveness, especially among residents at lower risk for severe COVID-19, who were generally tested less frequently and vaccinated later in the study period. Additional contributors may have included increasing bias in the composition of the unvaccinated group towards residents who declined vaccination, as well as cohort selection induced by heterogeneity in infection risk.^21^ For instance, if the vaccine offered partial (or “leaky”) protection,^22^ high infection risk within an unvaccinated group that is initially highly susceptible could induce selection bias over time as the most susceptible people are removed from the group, which would decrease estimated effectiveness of vaccination.

The study has several other limitations. First, our estimates of effectiveness focused on confirmed infections, not other important outcomes, such as symptomatic infections or severe disease. Incidence of hospitalizations and deaths in our cohort during the study period was too low to support rigorous analysis of those outcomes, and symptom reporting is unreliable in carceral settings.^23^ A related point is that we were only able to estimate effectiveness in relation to the date of test sample collection, not transmission date, which allows for the possibility that some detected infections might have preceded vaccination. Second, we evaluated effectiveness against any SARS-CoV-2 infection, not specific viral variants, because CDCR conducted limited viral genome sequencing during the study period. As the B.1.617.2 (delta) variant became dominant and cases rose in the general community over the months of June and July,^24,25^ CDCR detected a total of 286 cases among a population of nearly 99,000 residents during this period,^26^ a substantially lower rate when compared to the period between mid-March 2020 to mid-February 2021, during which the number of cases were above 200 in almost every week, peaking at 5,659 weekly cases in December 2020. Low incidence after February 2021 suggests that there may be substantial protection against outbreaks in this population with high levels of vaccination and prior infections, including during a period marked by increasing prevalence of more highly transmissible variants. However, as people continue to become infected and more outbreaks occur, further follow-up is necessary to reassess the effectiveness and protection afforded by vaccines. Third, CDCR used some antigen tests, which have lower sensitivity, potentially leading to under-detection of cases. However, at least 93% of all tests were PCR, so we expect any bias related to antigen testing to be minimal. Finally, the generalizability of our results to residents of jails and other correctional systems is unknown.

Residents of prisons and jails have borne a disproportionately large share of disease burden during the COVID-19 pandemic. Findings from this study–building on a growing evidence base indicating vaccine efficacy and effectiveness across a range of populations and settings–suggest that mRNA vaccines are extremely effective in protecting incarcerated people against infection, including residents at high risk of severe COVID-19 illness. Continued emphasis on vaccination and other ongoing mitigation practices are essential in preventing COVID-19 in this disproportionately affected population. Incarcerated people, correctional workers, and the wider community all stand to benefit from those efforts.

## Data Availability

Data not available due to Data Use Agreement between Stanford University and the California Department of Corrections and Rehabilitation.

## ACKNOWLEDGMENTS

We thank John Dunlap, Heidi Bauer and the other staff members at California Department of Corrections and Rehabilitation for providing data and assistance with interpretation of study results. We also acknowledge help from members of the Stanford-CIDE Coronavirus Simulation Model (SC-COSMO) consortium.

## Footnotes

* See e.g., 45 C.F.R. part 46, 21 C.F.R. part 56; 42 U.S.C. §241(d); 5 U.S.C. §552a; 44 U.S.C. §3501 et seq.

