## Supplemental Appendix for "Effectiveness of COVID-19 Vaccines among Incarcerated People in California State Prisons: A Retrospective Cohort Study"

**SUPPLEMENTARY APPENDIX**

*The findings and conclusions in this report are those of the authors and do not necessarily represent the official position of the Centers for Disease Control and Prevention.

1.	Cohort, study period and variables

#### 1.1 Construction of analytic cohort

Daily data extracts provided by the California Department of Corrections and Rehabilitation included a unique pseudo-identifier that allowed us to follow individual residents over time.

Residents were excluded from the analytic cohort if they:

- had confirmed infections prior to the start of the study period
- were newly incarcerated after the start of the study period
- had prior vaccination for SARS-CoV-2 (e.g. vaccinated in a jail or the community)
- had missing race or ethnic group, Covid-19 risk score, security level, or housing data

Figure S1 shows the application of these exclusion criteria and construction of the analytic cohort.

#### 1.2 Study period

The study period began on December 22, 2020, which was the day that CDCR’s vaccination program began.

We chose an end date of March 1, 2021 for the study period because case rates had declined by that date to a level of 3.8 per 10,000 person-days – a drop of approximately 95% compared to the start of the study period – and continued to decrease precipitously thereafter, to 0.5 per 10,000 by March 15 and 0.1 per 10,000 by July 1 (Figures S2 and S5). The period after March 1, 2021 was also marked by declining testing rates (from 818.5 tests per 10,000 person-days on March 1, 2021, to 554.3 by May 1, 2021 to 366.7 by July, 2021), and by a substantial fraction of the cohort having either received at least one vaccination dose or having tested positive.

#### 1.3 Variables

In addition to information on residents’ demographic characteristics, the data included various carceral characteristics with potential relevance for vaccine prioritization and exposure to infection risk—specifically security level, room type, and participation in penal labor. Table S4 describes the characteristics of the study population.

*Sex*. Male and female residents were generally housed in separate prisons, with 4% of the female incarcerated population housed in a prison that held both male and female residents. We did not adjust for sex given strong collinearity with prison fixed effects.

*Racial or ethnic group*. We grouped race and ethnicity information, assigned by CDCR based on a combination of self-reports and administrative records, into categories used by the U.S. Census Bureau (Hispanic or Latino, non-Hispanic Black or African American, non-Hispanic White, non-Hispanic American Indian or Alaska Native, non-Hispanic Asian or Pacific Islander, and non-Hispanic Other).^[1]^ The non-Hispanic Asian and non-Hispanic Hawaiian or other Pacific Islander groups were combined due to small sample size. Race or ethnic groupings were mutually exclusive.

*Covid-19 risk score*. The Covid-19 risk score was developed by CDCR to grade a resident’s risk of severe outcomes following SARS-CoV-2 infection. It is an index equal to the sum of weighted indicators for 17 items identified in the scientific literature as risk factors for severe Covid-19 outcomes (see Table S1 for details). For incorporation as a covariate in regression models on vaccine effectiveness, we top-coded the Covid-19 risk score to 4, which was classified as “high-risk” for all CDCR policies. For descriptive tables, we classified risk scores of 0-1 as “low”, scores of 2-3 “moderate”, and scores of 4 or greater “high”. Low-scoring residents were aged younger than 65 years and had no more than 1 comorbidity; moderate-scoring residents were aged younger than 65 years with 2 or 3 comorbidities; and high-scoring residents were aged 65 years and older or had 4 or more comorbidities.

*Prior SARS-CoV-2 infection*. The data included Covid-19 testing information. CDCR has undertaken extensive testing of residents for SARS-CoV-2 since April 2020, using real-time PCR and antigen tests (see Table S2 for details on testing programs for residents). We defined a prior SARS-CoV-2 infection as having had at least one positive test in CDCR’s clinical records prior to the beginning of the study period. Recent SARS-CoV-2 infections were defined as an initial infection within the 90 days prior to the beginning of the study period. Recent infections were excluded from the secondary analysis that included prior infections because of difficulties discerning positive tests from breakthrough infections or past infections. Table S5 describes Covid-19 cases among residents with prior infections.

*Security level*. CDCR rates each resident’s security level from 1 (lowest) to 4 (highest), based on a multifactorial assessment of the resident’s risk of misconduct; this rating influences housing placement and eligibility for activities such as visitations, recreation, and penal labor.

*Room type*. Residents are housed in rooms, discrete spaces that are at least partially enclosed by solid walls. We defined room type according to the number of residents housed in each room, dichotomizing this variable into cells (rooms with 1-2 occupants) and dormitories (rooms with 3 or more occupants).

*Penal labor*. Using information on residents’ involvement in work roles (e.g., janitorial, food preparation), we created a binary variable indicating whether each resident had participated in penal labor within the prior 14 days.

*Location*. CDCR recorded the location of residents at the end of each day. Location specified whether a resident was in CDCR custody, housed outside the CDCR system (jail, hospital), was released, or died while in custody. Residents with two or more days of missing location data were excluded from the analysis. Location information can be used to generate variables at different levels of granularity. The largest unit of observation was prison, followed by facility, building, housing unit, floor, and room. In our main regression analysis, we clustered standard errors at housing unit level, but we examined alternative levels of location units for defining clusters in secondary analyses.

*Staff case rate.* Information on confirmed infections among staff were available through routine mandatory testing for staff, separate from the resident testing programs described above and in Table S2. Staff were required to be tested at least once every 14 days, increased to once weekly at institutions reporting COVID-19 outbreaks. Using this information on infections among staff, we created a prison-day level variable of a 7-day rolling Covid-19 case rate among staff at each prison. The weekly rolling case rate represented the sum of cases from the past 7 days divided by the total number of active staff at the prison. The total number of staff were derived from monthly rosters of active staff members at each prison.

#### 1.4 Covid-19 outcomes and vaccination

The data included information about Covid-19 testing, hospitalizations, and deaths at the person-day record level.

*Confirmed infection.* A confirmed infection was defined as a positive real-time PCR or antigen diagnostic test for SARS-CoV-2. Positive tests were assigned to the date of specimen collection.

*Hospitalization.* We defined hospitalization related to a SARS-CoV-2 infection as a hospitalization that occurred within three days prior to or 14 days after an infection was initially confirmed. For purposes of attributing hospitalization events to person-days stratified by vaccination category (in Table 2 in the main text), the hospitalization event was associated with vaccination status at the time of sample collection for a confirmed infection.

*Death.* All deaths related to a SARS-CoV-2 infection were classified and confirmed by the California Correctional Health Care Services. For attribution to person-days stratified by vaccination category (in Table 2), deaths were assigned to the date of sample collection for a confirmed infection.

*Vaccination.* Prison healthcare staff recorded each dose offer, to whom it was made, and whether it was accepted or declined; this information was then entered into the CDCR electronic health record system. Residents who accepted a dose were vaccinated on the spot.

#### 1.5 Assignment of values to time-varying variables

Except for the race or ethnicity and sex variables, the variables in the CDCR person-day data were treated as time-varying in the analysis, meaning that their values changed for some residents during the study period.

### 2. Statistical analysis

#### 1.1 Kaplan-Meier survival curves

We estimated survival curves for the vaccinated and unvaccinated groups using the Kaplan-Meier method (Figure S6). In this unadjusted analysis, observation time for the vaccinated group began with receipt of a first vaccination dose. Observation time for the unvaccinated group came from those who were not vaccinated during the study period and from those who were vaccinated, prior to receiving their first dose. We estimated unadjusted survival curves from three starting points: the start of the vaccination program (December 22, 2020), when vaccination commenced in all prisons within the vaccinated cohort (January 21, 2021), and when over half of the vaccinated cohort received a first dose (February 2, 2021).

#### 1.2 Estimating vaccine effectiveness

We evaluated two different strategies for constructing the analytic cohort and estimating vaccine effectiveness. Based on key features of the vaccination campaign and dataset, our main analysis used a Cox proportional hazards regression model. Here we provide details on this approach, as well as describing the alternative approach that we considered, based on a rolling-entry matched cohort design.

##### 1.2.1. Survival analysis

We used the Andersen-Gill extension to the multivariable Cox proportional hazards regression model to estimate adjusted hazard ratios for confirmed SARS-CoV-2 infection associated with different categories of vaccination status. Observation time began at the start of the study period. Observation time ended on the last day a cohort member was in CDCR custody, at the day of sample collection of a confirmed infection, or at the end of the study period, whichever came first.

In an unadjusted analysis (reported in Table 2), the model included only indicator variables for vaccination status as explanatory variables, with no other covariates. Standard errors were clustered by housing unit to account for non-independence between persons.

Adjusted models controlled for residents’ race or ethnic group (Hispanic, non-Hispanic Black or African American, non-Hispanic White, non-Hispanic American Indian or Alaska Native, non-Hispanic Asian or Pacific Islander, non-Hispanic Other), Covid-19 risk score (0 to ≥4), security level (1, 2, 3, 4), room type (cell, dorm), involvement in penal labor (yes, no), and the prison-specific 7-day rolling Covid-19 case rate for staff (continuous). To account for non-independence between persons, we clustered standard errors by housing unit. Housing units are discrete cohorts within prisons, wherein residents participate in activities (e.g. recreation, laundry, prison commissary, dining) separate from other cohorts.

In addition, to help ensure unbiased estimates, we specified prison-level fixed effects, a modeling choice influenced by two considerations. First, unlike a random effects specification, fixed effects are more conservative in that they do not depend on an assumption of non-correlation with independent variables—an assumption that is likely violated in our analysis. Second, our study sample represented the majority of the CDCR population, and all CDCR prisons commenced vaccination during the study period.

In the main analysis, all residents were in custody at the start of the study period. The baseline hazard function accounted for differential risk of infection over the study period since calendar time was perfectly correlated with survival time.

We conducted several secondary analyses (Table S3), as follows:

A. Subgroup analyses:
   i. mRNA-1273 only: Because the majority of vaccines administered were mRNA-1273, we restricted vaccinated categories to include mRNA-1273 doses only. In this analysis, residents who received BNT162b2 doses were censored on the day before their first dose.
   ii. Medically-vulnerable residents: We re-estimated vaccine effectiveness on subsets of the study cohort having Covid-19 risk scores of at least 2, at least 3, or at least 4. Because CDCR includes age as a component in its Covid-19 risk score, all people 65 years and older had Covid-19 risk scores of 4 or higher.
B. Including prior confirmed infections and admissions: We allowed the inclusion of people with past confirmed infections and newly incarcerated people. We maintained the exclusion of those with recent infections (within the 90 days prior to the beginning of the study period) because of difficulties discerning positive tests from breakthrough infections or past infections. We added an indicator variable in the model for whether each resident had a prior confirmed infection. For all cohort members, observation time began on a person’s first day in CDCR custody or the start of the study period, whichever came last. Since calendar time was no longer perfectly correlated with survival time in this analysis, we controlled for varying risk of infection over calendar time in two ways: (1) using week-level fixed effects; (2) incorporating daily effects parameterized using a penalized-spline with 4 degrees of freedom (Figure S7).
C. Right censoring on last test date: We replaced end of the study period as a censoring event with collection date of cohort members’ last test. Thus, observation time ended on the earliest of the following: the last day of sample collection for a SARS-CoV-2 diagnostic test or the day of sample collection for a confirmed infection.
D. Clustering standard errors: We assessed the sensitivity of effectiveness estimates to the choice of location unit level at which we clustered standard errors. We re-estimated the results clustering standard errors at the prison, facility, building, housing unit, floor, room, and person level. All models except the prison model include prison-level fixed effects.
E. Modification of study period end date: The primary analysis was restricted to the first two and a half months of the vaccination program to capture the direct effects of vaccination during a period with ongoing infections and mass testing. After March 1, 2021, outbreaks subsided, mass testing became infrequent, and vaccination coverage was high; reductions in the risk of secondary transmission may have attenuated levels of effectiveness directly attributable to residents’ receipt of a vaccine. To examine how the net effects of these and other factors might affect vaccine effectiveness estimates over a longer programmatic period, we conducted analyses in which the study period end date was set to February 15, March 15, April 1, April 15, May 1, June 1, and July 1. A small number of Ad26.COV2.S (Janssen) vaccines were administered over the extended study period; between its introduction on March 10, 2021 and July 1, 2021, Ad26.COV2.S vaccines were administered to 0.5% of the study cohort. Given our focus on mRNA vaccines, we censored cohort members’ observation time on the day prior to receipt of a Ad26.COV2.S.

##### 1.2.2. Matched cohort design

An alternative study design—a rolling-entry matched cohort^[2]^—has potential to better address differences in time-varying characteristics between vaccinated and unvaccinated groups, and at least one previous study of Covid-19 vaccine effectiveness has used this design.^[3]^ We matched vaccine recipients and unvaccinated controls on variables with potential relevance for vaccine prioritization, exposure, or severity of Covid-19: race or ethnic group, security level, room type, Covid-19 risk score, and refusals. To control for differential risk of infection at the prison-level, we stratified matching by prison. The values of these variables for vaccine recipients were assigned by the date of the first dose. Newly vaccinated persons were matched to unvaccinated controls. If a newly vaccinated person had previously been selected as a control, both vaccinated and unvaccinated persons would be censored on the day prior to vaccination of the newly vaccinated person.

When matching newly vaccinated to unvaccinated persons on a 1:1 and 1:3 ratio, 18-51% of vaccinated persons were unmatched. When we allowed for replacements for unvaccinated matches, allowing unvaccinated controls to act as a comparison multiple times for cases vaccinated on the same day, ≤2% of vaccinated cases were unmatched. However, covariate balance was poor. All matching specifications had about 30-40 percentage-point higher levels of vaccine refusals by the end of the study period in the unvaccinated controls compared to the vaccinated cases. The mean number of follow-up days from the date an unvaccinated control was matched to their first vaccine offer was under 5 days.

Vaccine effectiveness estimates for each exposure category were defined as 1 minus the risk ratio for that category. The risk ratio represents a period-specific hazard over discretized periods of time post-vaccination relative to vaccinated cases. Risk was defined as the probability of having a confirmed infection within the period of interest. The risk ratio compared the risk of vaccinated cases to the risk of unvaccinated controls over a period of interest. We estimated vaccine effectiveness within 6 days of receiving a first dose. Estimates varied widely across matching specifications, ranging from 38 to 61% without replacement and -48 to 24% with replacement. The covariate imbalance of people who would later refuse vaccination is suggestive of substantial residual confounding, as was high vaccine effectiveness within 6 days of receiving a first dose in several of the matching specifications.

Given the rapid and non-randomized rollout of vaccination by prison, persons with similar baseline characteristics would likely be offered vaccination at the same time. These conditions bias the composition of the matched unvaccinated group towards persons with short follow-up periods and persons who refuse vaccination, who may face different risks of infection than those who accept vaccination. We thus pursued a Cox proportional hazards approach.

Analyses were performed using R software, version 3.5.2 (R Foundation for Statistical Computing).

### 3. Ethics approval

The study was approved by the institutional review board (IRB) at Stanford University (protocol #55835). The IRB approval of the study included a waiver of consent, on the basis that CDCR provided the Stanford research team with a limited data set without direct identifiers, the data had been collected for operational purposes, and the study could not practicably be carried out otherwise. It was reviewed by CDC and conducted according to applicable federal law and CDC policy.^[4]^

### 4. Supplementary tables and figures

#### Table S1. Covid-19 risk score components and weights

| Condition | Definition | Weighted Score |
| --- | --- | --- |
| Age 65+ | Chronologic age of 65 years or above | 4 |
| Advanced liver disease | Advanced liver disease (cirrhosis/end stage liver disease) | 2 |
| Asthma | Persistent asthma (moderate or severe) as defined by the  California Correctional Health Care Services (CCHCS) asthma condition specifications | 1 |
| Cancer | High risk cancer as defined by the CCHCS cancer condition specifications (excludes most diagnoses of skin cancer and “personal history of” cancers”) | 2 |
| Chronic Kidney Disease | Chronic kidney disease as defined by the CCHCS chronic kidney disease condition specifications | 1 |
| Advanced Chronic Kidney Disease/Renal Failure | Chronic kidney disease (Stage 5) as defined by the CCHCS Chronic Kidney Disease Condition Specifications OR currently receiving Hemodialysis | 1 |
| Chronic Lung Disease (other) | Cystic fibrosis, pneumoconiosis, or pulmonary fibrosis | 1 |
| COPD | Chronic obstructive pulmonary disease | 2 |
| Diabetes | Diabetes | 1 |
| Diabetes (high risk) | High risk diabetes as defined by the CCHCS diabetes condition specifications | 1 |
| Heart Disease | Any of the following cardiovascular disease conditions: Cerebrovascular, Congestive Heart Failure, Congenital Heart Disease, Ischemic Heart Disease, Peripheral Vascular Disease, Thromboembolic Disease, Valvular Disease, and Cardiovascular Disease‐Not Otherwise Specified. | 1 |
| Heart Disease (high risk) | High risk heart disease as defined by CCHCS condition specifications | 1 |
| Hemoglobin Disorder | Hemoglobin disorders as defined by CCHCS Condition Specifications for Hemoglobinopathy, including Sickle Cell Disorder. | 1 |
| HIV | HIV | 1 |
| HIV (poorly controlled) | HIV with a CD4 count < 200 | 1 |
| Hypertension | Hypertension as defined by the CCHCS Hypertension Condition Specifications | 1 |
| Immunocompromised | Any of the following conditions: aplastic anemia, histiocytosis, immunosuppressed, organ transplant, other transplant | 2 |
| Neurologic conditions | Dementia, Parkinson's Disease, Multiple Sclerosis, Myasthenia Gravis, or Neurologic Disorder as defined by CCHCS condition specifications | 1 |
| Obesity | Body mass index of 30 or above | 1 |
| Other high risk chronic condition | Any of the following conditions when they are high risk per CCHCS condition specifications: Coccidioidomycosis, Connective Tissue Disorder, Endocrine Disorder, or Vasculitis | 1 |
| Pregnant | Pregnant | 1 |

#### Table S2. Covid-19 testing strategies and definitions^[5]^

| Strategy | Individuals Tested | Goals of Testing | Timing and Frequency |
| --- | --- | --- | --- |
| Diagnostic testing | • Patients with symptoms consistent with COVID-19 | • Confirm diagnosis of COVID-19 | • Test when patient presents with symptoms |
| Outbreak response testing | • Exposed inmates and staff who are susceptible to infection (naïve or >90 days since first positive test) • Close contacts of a confirmed case of COVID-19 (e.g., housing, yard, worksite, shared ventilation) • Broad-based testing of asymptomatic inmates in the entire facility or institution | • Determine the extent of transmission in the institution • Identify patients who are infected for isolation and monitoring • Limit transmission with rapid isolation | • As soon as possible after one or more cases are identified among inmates or staff at an institution • Serial testing every 3-7 days of groups (e.g., housing, worksite) of exposed inmates who initially test negative until all tests are negative in a 14-day time frame |
| Quarantine Testing | • Asymptomatic close contacts of a confirmed case of COVID-19 (e.g., housing, yard, worksite) in quarantine • Other asymptomatic exposed inmates in quarantine | • Identify patients who are infected for isolation and monitoring • Identify inmate contacts eligible for release from quarantine | • At the beginning of quarantine (within 24 hours if not tested within last 7 days or at 3 days post-exposure), middle of quarantine, and end of quarantine (day 12-14) trial testing every 3-7 days of all susceptible inmate contacts who initially test negative until all tests are negative in a 14-day time frame |
| Risk-based routine testing | • Patients using aerosol-generating devices (e.g., nebulizers or continuous positive airway pressure [CPAP] devices) • Vulnerable patients (age >65, comorbidities, COVID-19 weighted risk score ≥3) | • Early diagnosis to reduce morbidity and mortality among patients at higher risk of severe COVID-19 | • At least monthly in facilities or institutions in non-outbreak settings (no new cases in 14 days) • Upon patient request |
| Inmate-worker routine testing | • Susceptible inmates working in enclosed spaces with staff or inmates outside of their housing unit, in healthcare or other areas where patients are isolated or quarantined for COVID-19, providing health aid or peer support, or in jobs that require movement about the facility or institution | • Identify infections in asymptomatic non-exposed inmate workers to prevent transmission to other inmates and staff | • Weekly testing offer for susceptible inmate workers in non-outbreak settings |
| Testing for movement | • Arrivals from a county jail, return from out-to-court • Inter-facility transfers, transfers to fire camps • Release to the community, parole, probation • Inter-facility transfers for mental health, medical, or dental services • Returns from medical/hospital, off-site appointments | • Improve the safety of transfers • Reduce the risk of introduction of the virus • Early identification of asymptomatic infection | Procedures for testing during movement vary. Details can be found in California Correctional Health Care Services Covid-19 Interim Guidance: Covid-19 screening and testing matrix for patient movement |
| Public health surveillance testing | • Susceptible individuals representing cohorts of patients that regularly comingle or share air space (e.g., housing unit, worksite), excluding quarantined patients | • Detect outbreaks in an early phase to limit spread and prevent morbidity and mortality • Needed for the transition to higher re-opening phase | • Weekly testing of a sample of susceptible patients from each identified cohort in non-outbreak settings • Test 25% (up to 25) patients per cohort each week |

#### Figure S1. Inclusion and exclusion criteria for the study population

In custody on December 22, 2020

62,743 persons / 3,633,372 person-days

Total in custody between

December 22, 2020 through March 1, 2021

98,595 persons / 5,922,337 person-days

Excluding recent infections

76,371 persons / 4,432,486 person-days

0 persons / 178 person-days were excluded after improper vaccine schedule (e.g. switch vaccine types, incorrect number of doses)

11 persons / 354 person-days were excluded for being vaccinated outside of CDCR

22,213 persons / 1,489,319 person-days were excluded for having recent confirmed infections

Excluding all prior confirmed infections

65,566 persons / 3,697,246 person-days

10,805 persons / 735,240 person-days were excluded for having prior confirmed infections

2,823 persons / 63,874 person-days were excluded for being admitted after December 22, 2020

21 persons / 1,470 person-days excluded for missing race or ethnic group data

1 person / 63 person-days excluded for missing Covid-19 risk scores

88 persons / 5,020 person-days excluded for missing security levels

1,926 persons / 129,349 person-days excluded for missing location data.

Main analysis, adjusted

60,707 persons / 3,497,470 person-days

#### Figure S2. Tests, confirmed infections and test positivity over the study period.

14-day rolling rates of testing, confirmed infections, and test positivity.^*^ Partially vaccinated status defined as ≥14 days after first dose until receipt of second dose. Fully vaccinated status defined as ≥14 days after a second dose.

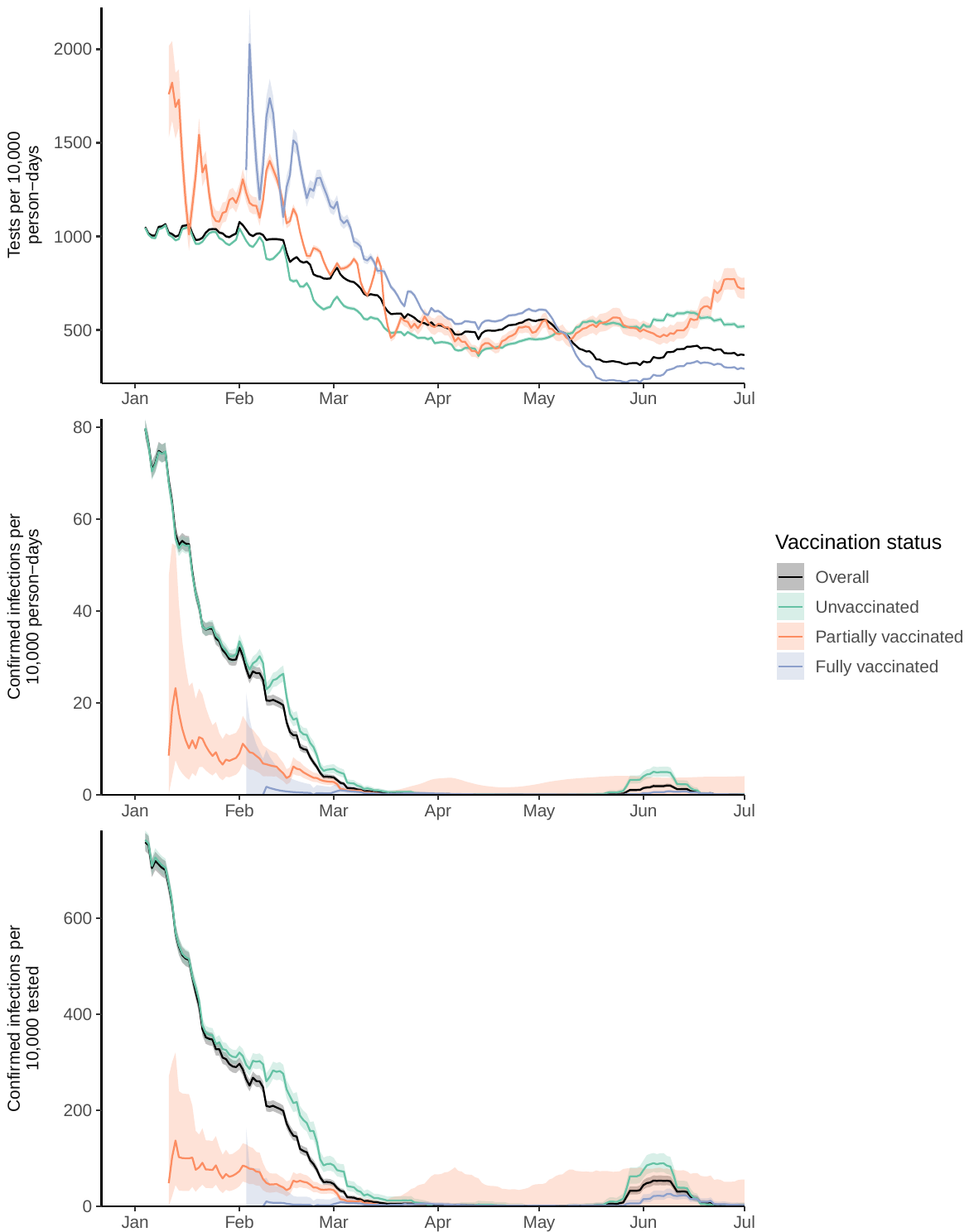

^*^Time periods with fewer than 200 people tested were excluded.

#### Figure S3. Vaccinations, testing and confirmed infections by prison

Each plot for a single prison shows cumulative percent vaccinated (black), 14-day rolling rates of tests per 10,000 person-days (purple) and confirmed infections (red) per 10,000 person-days over the period December 22, 2020 and July 1, 2021.* Shaded areas represent 95% confidence intervals. Denominators are based on prison populations as of December 22, 2020. Prison abbreviations listed in Table S6.

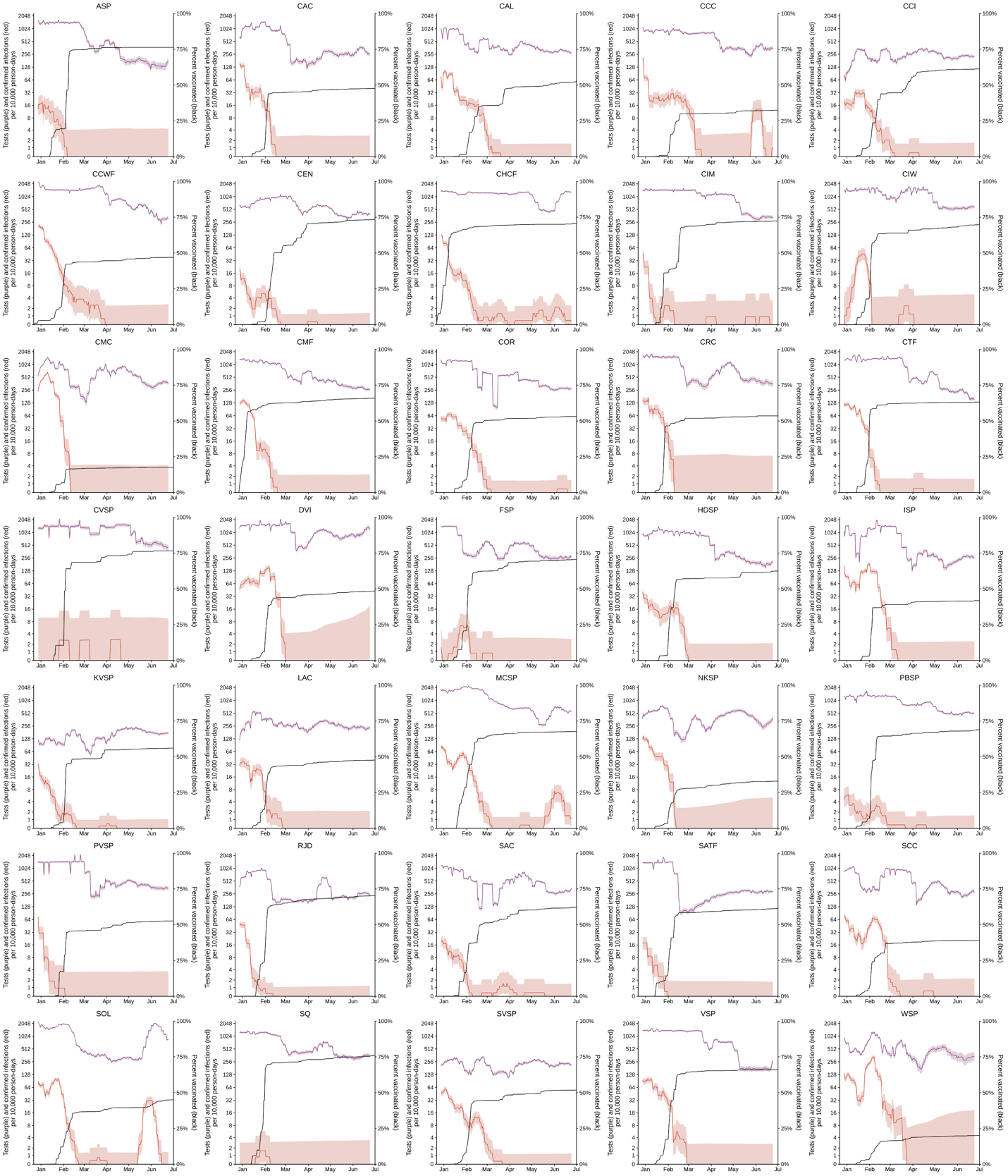

*Time periods with fewer than 20 people tested were excluded.

#### Figure S4. Testing by vaccination status and prison

14-day rolling rates of tests per 10,000 person-days, by vaccination category, stratified by prison over the period December 22, 2020 and July 1, 2021.* Shaded areas represent 95% confidence intervals. Partially vaccinated status defined as ≥14 days after a first dose until receipt of a second dose; fully vaccinated status defined as ≥14 days after a second dose. Prison abbreviations listed in Table S6.

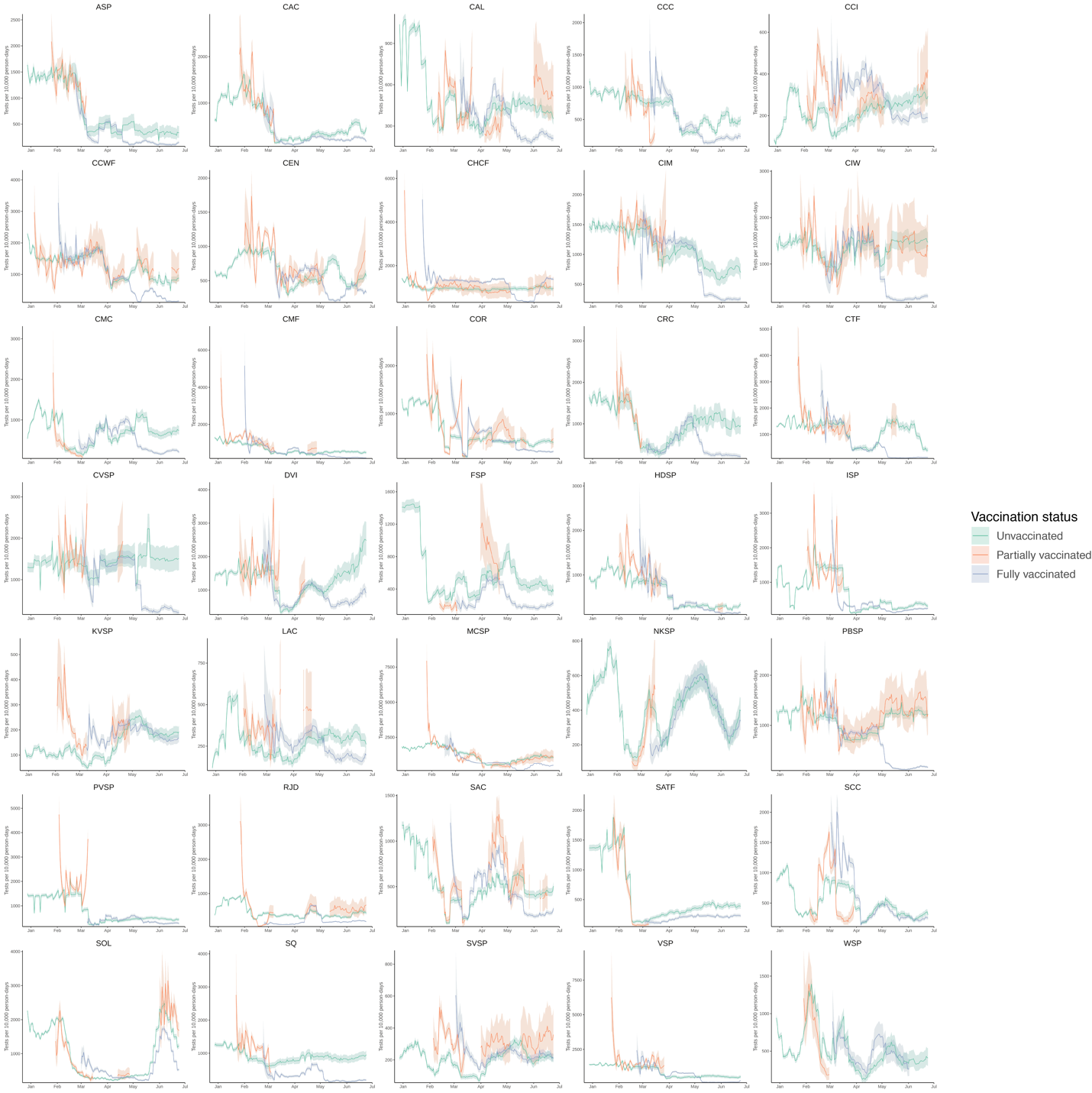

*Time periods with fewer than 20 people tested were excluded.

#### Table S3. Results from secondary analyses

A. Estimated vaccine effectiveness in specific subgroups of interest.

|  | **Including mRNA-1273 (Moderna) only** | **Including Covid-19 risk scores* ≥2 only** | **Including Covid-19 risk scores* ≥3 only** | **Including Covid-19 risk scores* ≥4 only** |
| --- | --- | --- | --- | --- |
| **Covid-19 vaccination status** | *Estimate (95% CI)* | *Estimate (95% CI)* | *Estimate (95% CI)* | *Estimate (95% CI)* |
| *Unvaccinated* | *Ref.* | *Ref.* | *Ref.* | *Ref.* |
| *Vaccinated with one dose* |  |  |  |  |
| 0-6d after first dose | -8% (-47 to 20) | 12% (-24 to 38) | 19% (-19 to 45) | 16% (-28 to 45) |
| 7-13d after first dose | 28% (-2 to 50) | 36% (2 to 58) | 39% (2 to 62) | 39% (-1 to 63) |
| ≥14d after first dose until second dose | 71% (58 to 80) | 74% (62 to 82) | 72% (56 to 81) | 73% (55 to 84) |
| *Vaccinated with two doses* |  |  |  |  |
| 0-13d after second dose | 86% (42 to 97) | 76% (40 to 90) | 74% (33 to 90) | 67% (8 to 88) |
| ≥14d after second dose | 96% (67 to 99) | 92% (74 to 98) | 94% (75 to 98) | 90% (61 to 97) |

^*^See Supplementary Materials Table S1

B. Estimated vaccine effectiveness in an expanded cohort including people with prior infections and people newly incarcerated during the study period.

|  | **Unadjusted** | **Adjusted, weekly fixed effects**^†^ | **Adjusted, daily spline**^†^ |
| --- | --- | --- | --- |
| **Covid-19 vaccination status** | *Estimate (95% CI)* | *Estimate (95% CI)* | *Estimate (95% CI)* |
| *Unvaccinated* | *Ref.* | *Ref.* | *Ref.* |
| *Vaccinated with one dose* |  |  |  |
| 0-6d after first dose | -27% (-74 to 8) | 16% (-14 to 39) | 16% (-14 to 39) |
| 7-13d after first dose | 7% (-35 to 36) | 43% (19 to 60) | 43% (19 to 60) |
| ≥14d after first dose until second dose | 52% (33 to 66) | 74% (63 to 81) | 74% (63 to 81) |
| *Vaccinated with two doses* |  |  |  |
| 0-13d after second dose | 67% (24 to 86) | 85% (66 to 94) | 85% (66 to 94) |
| ≥14d after second dose | 90% (67 to 97) | 96% (88 to 99) | 96% (88 to 99) |

^†^We controlled for calendar time using week-level fixed effects and by incorporating daily effects parameterized using a penalized-spline with 4 degrees of freedom.

C. Estimated vaccine effectiveness with censoring of observation time on date of collection of last test.

|  | **Right censor**  **by testing** |
| --- | --- |
| **Covid-19 vaccination status** | *Estimate (95% CI)* |
| *Unvaccinated* | *Ref.* |
| *Vaccinated with one dose* |  |
| 0-6d after first dose | 21% (-8 to 42) |
| 7-13d after first dose | 48% (26 to 64) |
| ≥14d after first dose until second dose | 77% (68 to 84) |
| *Vaccinated with two doses* |  |
| 0-13d after second dose | 88% (72 to 95) |
| ≥14d after second dose | 98% (91 to 99) |

D. Estimated vaccine effectiveness with standard errors clustered at different levels.

|  | **Level for clustering of standard errors** | | | | | | |
| --- | --- | --- | --- | --- | --- | --- | --- |
|  | **Prison** | **Facility** | **Building** | **Housing unit**  **(*main analysis*)** | **Floor** | **Room** | **Person** |
| **Covid-19 vaccination status** | *Estimate (95% CI)* | *Estimate (95% CI)* | *Estimate (95% CI)* | *Estimate (95% CI)* | *Estimate (95% CI)* | *Estimate (95% CI)* | *Estimate (95% CI)* |
| *Unvaccinated* | *Ref.* | *Ref.* | *Ref.* | *Ref.* | *Ref.* | *Ref.* | *Ref.* |
| *Vaccinated with one dose* |  |  |  |  |  |  |  |
| 0-6d after first dose | 12% (-49 to 48) | 16% (-29 to 46) | 16% (-14 to 39) | 16% (-15 to 39) | 16% (-7 to 35) | 16% (4 to 27) | 16% (8 to 24) |
| 7-13d after first dose | 41% (-26 to 73) | 44% (1 to 68) | 44% (19 to 61) | 44% (20 to 61) | 44% (24 to 59) | 44% (32 to 54) | 44% (36 to 51) |
| ≥14d after first dose until second dose | 72% (57 to 81) | 74% (62 to 83) | 74% (63 to 82) | 74% (64 to 82) | 74% (65 to 81) | 74% (68 to 79) | 74% (68 to 79) |
| *Vaccinated with two doses* |  |  |  |  |  |  |  |
| 0-13d after second dose | 81% (63 to 90) | 85% (61 to 95) | 85% (66 to 94) | 85% (66 to 94) | 85% (68 to 93) | 85% (71 to 93) | 85% (79 to 90) |
| ≥14d after second dose | 95% (87 to 98) | 97% (90 to 99) | 97% (88 to 99) | 97% (88 to 99) | 97% (88 to 99) | 97% (89 to 99) | 97% (89 to 99) |

E. Estimated vaccine effectiveness using alternative study end dates.

Study cohort characteristics for the modified end dates included 14-day rolling rates of testing and cases by vaccination status, and cumulative cases. All study cohort characteristics were calculated relative to the end date.

|  | **Modification of study endpoint** | | | | | | | |
| --- | --- | --- | --- | --- | --- | --- | --- | --- |
|  | **Feb 15, 2021** | **Mar 1, 2021** | **Mar 15, 2021** | **Apr 1, 2021** | **Apr 15, 2021** | **May 1, 2021** | **June 1, 2021** | **July 1, 2021** |
| **Covid-19 vaccination status** | *Estimate*  *(95% CI)* | *Estimate*  *(95% CI)* | *Estimate*  *(95% CI)* | *Estimate (95% CI)* | *Estimate (95% CI)* | *Estimate (95% CI)* | *Estimate (95% CI)* | *Estimate (95% CI)* |
| *Unvaccinated* | *Ref.* | *Ref.* | *Ref.* | *Ref.* | *Ref.* | *Ref.* | *Ref.* | *Ref.* |
| *Vaccinated with one dose* |  |  |  |  |  |  |  |  |
| 0-6d after first dose | 17% (-14 to 40) | 16% (-15 to 39) | 16% (-15 to 39) | 16% (-15 to 38) | 16% (-16 to 38) | 15% (-16 to 38) | 15% (-16 to 38) | 15% (-16 to 38) |
| 7-13d after first dose | 47% (21 to 64) | 44% (20 to 61) | 44% (19 to 61) | 43% (19 to 60) | 43% (19 to 60) | 43% (19 to 60) | 43% (19 to 60) | 43% (19 to 60) |
| ≥14d after first dose until second dose | 83% (76 to 89) | 74% (64 to 82) | 75% (64 to 82) | 74% (64 to 82) | 74% (64 to 82) | 74% (64 to 82) | 74% (64 to 82) | 74% (64 to 82) |
| *Vaccinated with two doses* |  |  |  |  |  |  |  |  |
| 0-13d after second dose | 81% (53 to 92) | 85% (66 to 94) | 84% (67 to 92) | 84% (67 to 92) | 84% (67 to 92) | 84% (67 to 92) | 84% (67 to 92) | 84% (67 to 92) |
| ≥14d after second dose | 98% (82 to 100) | 97% (88 to 99) | 94% (85 to 98) | 84% (71 to 92) | 83% (69 to 90) | 81% (66 to 89) | 82% (69 to 89) | 82% (69 to 89) |
| **Study cohort characteristics** |  |  |  |  |  |  |  |  |
| *Testing rates*^*^ |  |  |  |  |  |  |  |  |
| Overall | 919.4 (912.0 to 926.8) | 818.5 (811.4 to 825.5) | 672.1 (665.7 to 678.5) | 529.7 (524.0 to 535.5) | 497.5 (491.9 to 503.1) | 554.3 (548.4 to 560.3) | 332.7 (328.1 to 337.4) | 366.7 (361.8 to 371.7) |
| Unvaccinated | 879.0 (868.7 to 889.3) | 664.6 (654.3 to 675.0) | 545.0 (535.5 to 554.7) | 433.1 (424.3 to 442.1) | 397.4 (388.6 to 406.4) | 450.3 (440.7 to 460.0) | 529.5 (518.8 to 540.4) | 519.6 (508.5 to 530.8) |
| ≥14d after first dose until second dose | 1075.3 (1048.7 to 1102.4) | 830.3 (817.8 to 843.0) | 848.3 (826.3 to 870.6) | 540.4 (496.4 to 587.4) | 432.5 (403.2 to 463.3) | 522.3 (486.6 to 559.9) | 503.6 (459.1 to 551.2) | 722.1 (667.9 to 779.5) |
| ≥14d after second dose | 1284.2 (1225.0 to 1345.4) | 1164.0 (1128.4 to 1200.6) | 820.5 (804.2 to 837.0) | 608.0 (599.9 to 616.3) | 552.6 (545.1 to 560.1) | 609.3 (601.5 to 617.2) | 239.0 (234.2 to 243.8) | 292.8 (287.6 to 298.2) |
| *Case rates*^*^ |  |  |  |  |  |  |  |  |
| Overall | 15.9 (14.9 to 16.9) | 3.8 (3.4 to 4.3) | 0.5 (0.3 to 0.7) | 0.2 (0.1 to 0.3) | 0.1 (0.1 to 0.3) | 0.1 (0.0 to 0.2) | 1.5 (1.2 to 1.8) | 0.1 (0.0 to 0.2) |
| Unvaccinated | 21.5 (19.9 to 23.1) | 5.7 (4.8 to 6.7) | 0.9 (0.5 to 1.3) | 0.0 (0.0 to 0.2) | 0.2 (0.0 to 0.5) | 0.1 (0.0 to 0.3) | 4.0 (3.1 to 5.1) | 0.1 (0.0 to 0.4) |
| ≥14d after first dose until second dose | 3.6 (2.3 to 5.6) | 2.8 (2.1 to 3.6) | 0.1 (0.0 to 0.8) | 0.0 (0.0 to 3.6) | 0.0 (0.0 to 2.0) | 0.0 (0.0 to 2.4) | 0.0 (0.0 to 4.0) | 0.0 (0.0 to 4.1) |
| ≥14d after second dose | 0.7 (0.0 to 4.0) | 0.6 (0.1 to 2.1) | 0.3 (0.1 to 0.7) | 0.3 (0.1 to 0.5) | 0.1 (0.0 to 0.3) | 0.1 (0.0 to 0.3) | 0.4 (0.2 to 0.6) | 0.1 (0.0 to 0.2) |
| *Cumulative cases (%)* | 21.37% | 21.77% | 21.82% | 21.85% | 21.86% | 21.87% | 22.02% | 22.11% |

^*^Per 10,000 person-days

#### Table S4. Demographic, clinical, and carceral characteristics stratified by counts and person-days

|  | Persons (N=60,707) | Vaccinated (N=29,947) | Unvaccinated (Person-days=2,633,734) | 0-6 days after first dose (Person-days=206,960) | 7-13 days after first dose (Person-days=199,746) | ≥14 days after first dose (Person-days=286,856) | 0-13 days after second dose (Person-days=120,141) | ≥14 days after second dose (Person-days=50,033) |
| --- | --- | --- | --- | --- | --- | --- | --- | --- |
| Demographic Characteristics | | | | | | | | |
| Age category - no. (%) | | | | |  |  |  |  |
| 18-39y | 29,922 (49.3%) | 12,378 (41.3%) | 1,450,103 (55.1%) | 85,557 (41.3%) | 82,170 (41.1%) | 112,091 (39.1%) | 23,580 (19.6%) | 8,301 (16.6%) |
| 40-59y | 23,469 (38.7%) | 12,888 (43%) | 968,631 (36.8%) | 89,248 (43.1%) | 86,613 (43.4%) | 124,154 (43.3%) | 54,410 (45.3%) | 19,904 (39.8%) |
| ≥60y | 7,316 (12.1%) | 4,681 (15.6%) | 215,000 (8.2%) | 32,155 (15.5%) | 30,963 (15.5%) | 50,611 (17.6%) | 42,151 (35.1%) | 21,828 (43.6%) |
| Race or ethnicity^*^ - no. (%) | | | | | | | | |
| Hispanic or Latino | 25,914 (42.7%) | 13,459 (44.9%) | 1,131,603 (43.0%) | 93,065 (45.0%) | 89,854 (45.0%) | 126,905 (44.2%) | 37,734 (31.4%) | 11,965 (23.9%) |
| Black or African American | 19,894 (32.8%) | 8,166 (27.3%) | 925,772 (35.2%) | 56,375 (27.2%) | 54,203 (27.1%) | 77,923 (27.2%) | 39,082 (32.5%) | 17,682 (35.3%) |
| White | 10,957 (18.0%) | 6,247 (20.9%) | 414,974 (15.8%) | 43,183 (20.9%) | 41,817 (20.9%) | 61,856 (21.6%) | 33,894 (28.2%) | 16,370 (32.7%) |
| American Indian or Alaska Native | 670 (1.1%) | 325 (1.1%) | 27,083 (1.0%) | 2,239 (1.1%) | 2,190 (1.1%) | 3,213 (1.1%) | 1,542 (1.3%) | 726 (1.5%) |
| Asian or Pacific Islander | 833 (1.4%) | 422 (1.4%) | 36,139 (1.4%) | 2,911 (1.4%) | 2,768 (1.4%) | 4,057 (1.4%) | 1,620 (1.3%) | 552 (1.1%) |
| Other | 2,439 (4.0%) | 1,328 (4.4%) | 98,163 (3.7%) | 9,187 (4.4%) | 8,914 (4.5%) | 12,902 (4.5%) | 6,269 (5.2%) | 2,738 (5.5%) |
| Sex - no. (%) | | | | | | | | |
| Male | 58,017 (95.6%) | 28,636 (95.6%) | 2,533,272 (96.2%) | 197,821 (95.6%) | 190,761 (95.5%) | 270,745 (94.4%) | 114,693 (95.5%) | 48,516 (97.0%) |
| Female | 2,661 (4.4%) | 1,311 (4.4%) | 100,433 (3.8%) | 9,139 (4.4%) | 8,985 (4.5%) | 16,111 (5.6%) | 5,448 (4.5%) | 1,517 (3.0%) |
| Clinical Characteristics | | | | | | | | |
| Covid-19 risk score category^†^ - no. (%) | | | | | | | | |
| Low (0-1) | 42,093 (69.3%) | 18,829 (62.9%) | 1,976,044 (75.0%) | 130,198 (62.9%) | 125,366 (62.8%) | 169,569 (59.1%) | 37,938 (31.6%) | 13,574 (27.1%) |
| Moderate (2-3) | 11,509 (19.0%) | 6,415 (21.4%) | 459,034 (17.4%) | 44,447 (21.5%) | 43,290 (21.7%) | 63,957 (22.3%) | 33,371 (27.8%) | 11,524 (23.0%) |
| High (≥4) | 7,105 (11.7%) | 4,703 (15.7%) | 198,656 (7.5%) | 32,315 (15.6%) | 31,090 (15.6%) | 53,330 (18.6%) | 48,832 (40.6%) | 24,935 (49.8%) |
| Medical conditions - no. (%) | | | | | | | | |
| Any pre-existing condition^‡^ | 51,129 (84.2%) | 25,881 (86.4%) | 2,166,816 (82.3%) | 178,862 (86.4%) | 172,762 (86.5%) | 250,530 (87.3%) | 112,216 (93.4%) | 47,236 (94.4%) |
| Any immunocompromising condition^§^ | 2,031 (3.3%) | 1,349 (4.5%) | 64,792 (2.5%) | 9,289 (4.5%) | 8,921 (4.5%) | 14,518 (5.1%) | 12,498 (10.4%) | 6,222 (12.4%) |
| Advanced Liver Disease | 2,141 (3.5%) | 1,454 (4.9%) | 64,178 (2.4%) | 10,045 (4.9%) | 9,746 (4.9%) | 15,495 (5.4%) | 13,578 (11.3%) | 6,593 (13.2%) |
| Asthma | 8,307 (13.7%) | 4,049 (13.5%) | 354,568 (13.5%) | 28,005 (13.5%) | 27,101 (13.6%) | 40,260 (14.0%) | 19,740 (16.4%) | 8,537 (17.1%) |
| Cancer | 1,773 (2.9%) | 1,159 (3.9%) | 54,810 (2.1%) | 7,980 (3.9%) | 7,661 (3.8%) | 12,391 (4.3%) | 10,580 (8.8%) | 5,238 (10.5%) |
| Chronic Kidney Disease | 8,889 (14.6%) | 5,406 (18.1%) | 308,404 (11.7%) | 37,313 (18.0%) | 36,136 (18.1%) | 57,150 (19.9%) | 40,756 (33.9%) | 18,090 (36.2%) |
| Coccidioidomycosis | 522 (0.9%) | 269 (0.9%) | 21,332 (0.8%) | 1,858 (0.9%) | 1,813 (0.9%) | 2,798 (1.0%) | 1,523 (1.3%) | 704 (1.4%) |
| Chronic Obstructive Pulmonary Disease | 1,757 (2.9%) | 1,207 (4.0%) | 47,752 (1.8%) | 8,281 (4.0%) | 7,981 (4.0%) | 13,036 (4.5%) | 12,217 (10.2%) | 7,751 (15.5%) |
| Connective Tissue Disorder | 481 (0.8%) | 314 (1.0%) | 16,607 (0.6%) | 2,173 (1.0%) | 2,127 (1.1%) | 3,418 (1.2%) | 2,764 (2.3%) | 1,326 (2.7%) |
| Cardiovascular Disease | 3,115 (5.1%) | 1,943 (6.5%) | 101,882 (3.9%) | 13,421 (6.5%) | 13,035 (6.5%) | 20,294 (7.1%) | 16,990 (14.1%) | 11,748 (23.5%) |
| Dementia or Parkinson's | 432 (0.7%) | 300 (1.0%) | 11,807 (0.4%) | 2,067 (1.0%) | 1,989 (1.0%) | 3,030 (1.1%) | 3,176 (2.6%) | 3,382 (6.8%) |
| Diabetes | 4,886 (8.0%) | 3,090 (10.3%) | 152,451 (5.8%) | 21,247 (10.3%) | 20,513 (10.3%) | 33,250 (11.6%) | 26,746 (22.3%) | 13,698 (27.4%) |
| On Dialysis | 59 (0.1%) | 41 (0.1%) | 827 (0.0%) | 287 (0.1%) | 284 (0.1%) | 312 (0.1%) | 497 (0.4%) | 765 (1.5%) |
| Endocrine Disorder | 80 (0.1%) | 54 (0.2%) | 2,027 (0.1%) | 372 (0.2%) | 368 (0.2%) | 570 (0.2%) | 550 (0.5%) | 433 (0.9%) |
| Hemoglobinopathy | 648 (1.1%) | 326 (1.1%) | 26,979 (1.0%) | 2,258 (1.1%) | 2,191 (1.1%) | 3,477 (1.2%) | 2,046 (1.7%) | 761 (1.5%) |
| HIV | 481 (0.8%) | 309 (1.0%) | 17,317 (0.7%) | 2,110 (1.0%) | 2,000 (1.0%) | 2,695 (0.9%) | 2,392 (2.0%) | 1,064 (2.1%) |
| Hypertension | 15,068 (24.8%) | 8,786 (29.3%) | 546,879 (20.8%) | 60,674 (29.3%) | 58,908 (29.5%) | 92,366 (32.2%) | 62,425 (52.0%) | 28,515 (57.0%) |
| Immunocompromised | 844 (1.4%) | 564 (1.9%) | 27,997 (1.1%) | 3,862 (1.9%) | 3,709 (1.9%) | 6,029 (2.1%) | 5,086 (4.2%) | 2,768 (5.5%) |
| Lung Disease | 99 (0.2%) | 62 (0.2%) | 2,574 (0.1%) | 427 (0.2%) | 415 (0.2%) | 701 (0.2%) | 668 (0.6%) | 432 (0.9%) |
| Multiple Sclerosis | 41 (0.1%) | 28 (0.1%) | 1,519 (0.1%) | 194 (0.1%) | 189 (0.1%) | 247 (0.1%) | 264 (0.2%) | 232 (0.5%) |
| Myasthenia Gravis | 21 (0.0%) | 11 (0.0%) | 836 (0.0%) | 77 (0.0%) | 77 (0.0%) | 103 (0.0%) | 91 (0.1%) | 85 (0.2%) |
| Neurologic Disorder | 123 (0.2%) | 80 (0.3%) | 3,575 (0.1%) | 541 (0.3%) | 516 (0.3%) | 695 (0.2%) | 815 (0.7%) | 973 (1.9%) |
| Pregnancy | N < 10 | | | | | | | |
| Vasculitis | 24 (0.0%) | 16 (0.1%) | 793 (0.0%) | 112 (0.1%) | 112 (0.1%) | 201 (0.1%) | 188 (0.2%) | 102 (0.2%) |
| Overweight^‖^ | 21,137 (34.8%) | 10,173 (34.0%) | 932,901 (35.4%) | 70,268 (34%) | 67,633 (33.9%) | 95,093 (33.2%) | 34,811 (29.0%) | 13,337 (26.7%) |
| Obesity^‖^ | 21,960 (36.2%) | 11,386 (38.0%) | 913,646 (34.7%) | 78,732 (38.0%) | 76,241 (38.2%) | 112,202 (39.1%) | 51,920 (43.2%) | 19,599 (39.2%) |
| Severe Obesity^‖^ | 2,553 (4.2%) | 1,414 (4.7%) | 100,148 (3.8%) | 9,782 (4.7%) | 9,500 (4.8%) | 14,582 (5.1%) | 7,915 (6.6%) | 3,121 (6.2%) |
| Disability - no. (%) |  |  |  |  |  |  |  |  |
| Any disability^¶^ | 23,422 (38.6%) | 12,892 (43.0%) | 939,066 (35.7%) | 89,150 (43.1%) | 86,473 (43.3%) | 124,797 (43.5%) | 76,531 (63.7%) | 42,585 (85.1%) |
| Cognitive | 993 (1.6%) | 664 (2.2%) | 34,761 (1.3%) | 4,559 (2.2%) | 4,344 (2.2%) | 6,084 (2.1%) | 4,900 (4.1%) | 4,506 (9.0%) |
| Hearing | 2,033 (3.3%) | 1,319 (4.4%) | 65,679 (2.5%) | 9,092 (4.4%) | 8,786 (4.4%) | 13,776 (4.8%) | 10,732 (8.9%) | 6,168 (12.3%) |
| Mental Health | 19,467 (32.1%) | 10,510 (35.1%) | 810,241 (30.8%) | 72,731 (35.1%) | 70,696 (35.4%) | 100,252 (34.9%) | 57,864 (48.2%) | 31,187 (62.3%) |
| Mobility | 6,980 (11.5%) | 4,453 (14.9%) | 225,887 (8.6%) | 30,734 (14.9%) | 29,728 (14.9%) | 45,170 (15.7%) | 38,177 (31.8%) | 25,945 (51.9%) |
| Speech | 96 (0.2%) | 71 (0.2%) | 2,614 (0.1%) | 497 (0.2%) | 469 (0.2%) | 615 (0.2%) | 710 (0.6%) | 780 (1.6%) |
| Vision | 495 (0.8%) | 323 (1.1%) | 16,273 (0.6%) | 2,223 (1.1%) | 2,133 (1.1%) | 3,181 (1.1%) | 2,881 (2.4%) | 2,473 (4.9%) |
| Carceral Characteristics | | | | | | | | |
| Room type - no. (%) | | | | | | | | |
| Cell | 45,304 (74.6%) | 22,954 (76.6%) | 2,083,226 (79.1%) | 158,825 (76.7%) | 154,220 (77.2%) | 222,057 (77.4%) | 84,885 (70.7%) | 38,323 (76.6%) |
| Dorm | 15,403 (25.4%) | 6,993 (23.4%) | 550,508 (20.9%) | 48,135 (23.3%) | 45,526 (22.8%) | 64,799 (22.6%) | 35,256 (29.3%) | 11,710 (23.4%) |
| Security level - no. (%) | | | | | | | | |
| 1 (minimum) | 4,953 (8.2%) | 2,041 (6.8%) | 205,079 (7.8%) | 13,994 (6.8%) | 12,827 (6.4%) | 17,654 (6.2%) | 7,004 (5.8%) | 2,290 (4.6%) |
| 2 | 24,729 (40.7%) | 13,247 (44.2%) | 895,129 (34.0%) | 91,442 (44.2%) | 88,091 (44.1%) | 128,737 (44.9%) | 74,131 (61.7%) | 33,336 (66.6%) |
| 3 | 10,763 (17.7%) | 4,884 (16.3%) | 510,895 (19.4%) | 33,677 (16.3%) | 32,525 (16.3%) | 48,145 (16.8%) | 12,917 (10.8%) | 4,102 (8.2%) |
| 4 (maximum) | 20,262 (33.4%) | 9,775 (32.6%) | 1,022,631 (38.8%) | 67,847 (32.8%) | 66,303 (33.2%) | 92,320 (32.2%) | 26,089 (21.7%) | 10,305 (20.6%) |
| Participation in penal labor - no. (%) | 15,153 (25.0%) | 7,478 (25.0%) | 637,255 (24.2%) | 51,672 (25.0%) | 49,534 (24.8%) | 73,719 (25.7%) | 29,753 (24.8%) | 10,111 (20.2%) |

^*^All categories other than “Hispanic or Latino” are categorized as non-Hispanic ethnicity.

^†^Based on CDCR risk score. See Supplementary Materials Table S1.

^‡^Refers to the set of conditions identified by the Centers for Disease Control and Prevention as risk factors for increased risk of severe Covid-19 illness among adults of any age, specifically: advanced liver disease, asthma, cancer, chronic kidney disease, chronic obstructive pulmonary disease, cardiovascular disease, dementia, Parkinson’s, diabetes, on dialysis, hemoglobinopathy disorders, HIV, hypertension, immunocompromised, lung disease, neurologic disorders, pregnancy, vasculitis, overweight, obesity, and severe obesity.

^§^Refers to diagnosis of immunocompromised, severe HIV, or severe cancer.

^‖^Overweight refers to 25 < BMI < 30; obesity refers to 30 ≤ BMI < 40; severe obesity refers to 40 ≤ BMI.

^¶^Refers to presence of disability in six categories: cognitive, hearing, mental health, mobility, speech, and vision.

#### Figure S5. Cumulative vaccinations, infections, and releases.

Cumulative vaccinations with one or two doses of mRNA vaccines (top panel) and cumulative confirmed infections and releases from incarceration (bottom panel).

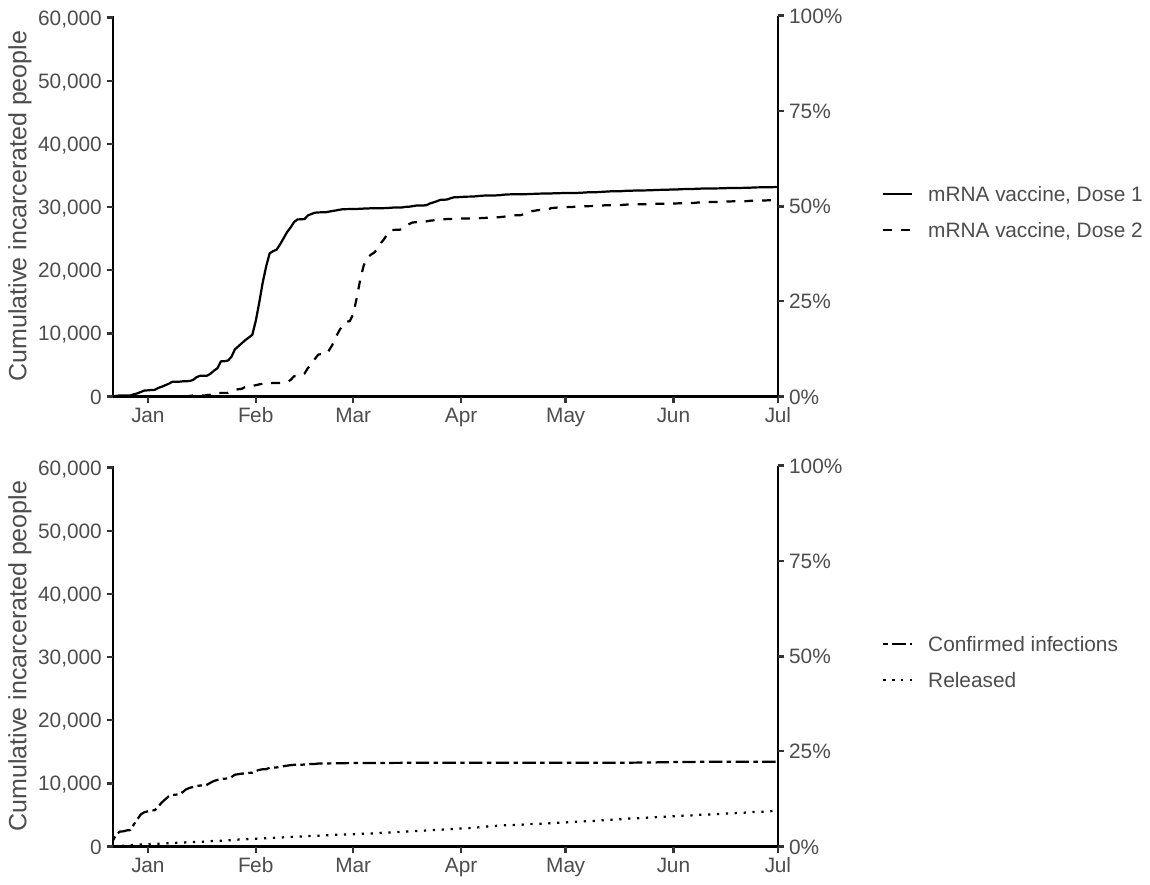

#### Figure S6. Unadjusted survival curves.

Kaplan-Meier curves of cumulative incidence of confirmed infections among unvaccinated and vaccinated persons.

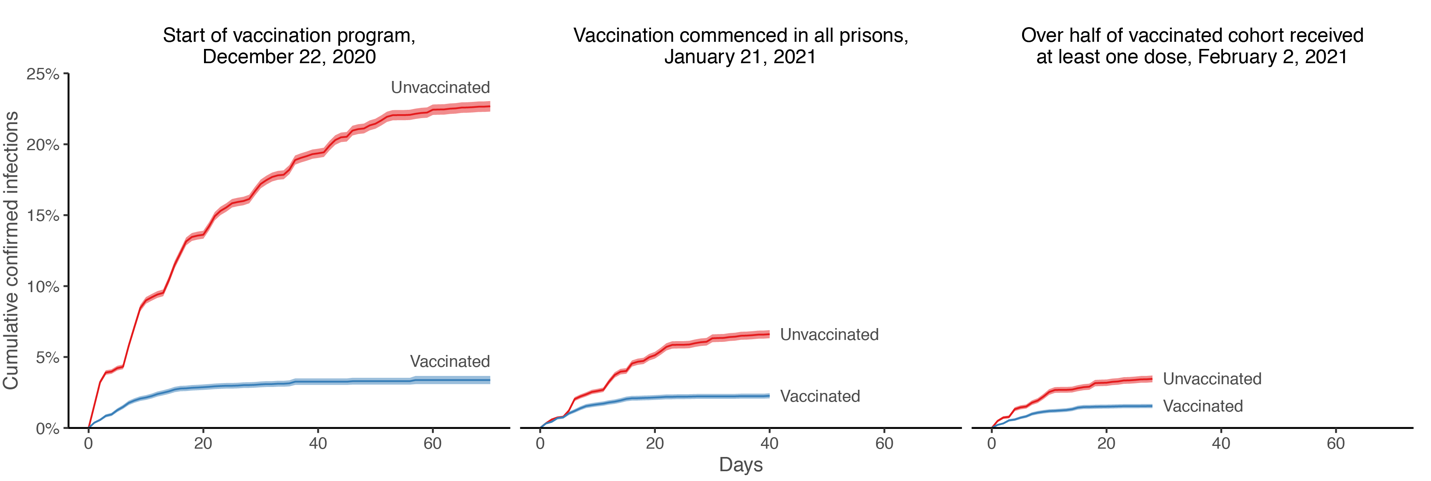

#### Table S5. Counts among study population with prior confirmed infections

| **Covid-19 vaccination status** | **Positive** | **Hospitalized^*^** | **Died^†^** | **Tested** | **Total** | **Median follow-up, days** | **Person-days** | **Positive per 10,000 person-days** |
| --- | --- | --- | --- | --- | --- | --- | --- | --- |
| **Unvaccinated** | 82 | 5 | 0 | 6,249 | 10,785 | 70 | 684,602 | 1.2 |
| **Vaccinated with one dose** |  |  |  |  |  |  |  |  |
| 0-6 days after first dose | 1 | 0 | 0 | 1,044 | 2,740 | 7 | 18,161 | 0.6 |
| 7-13 days after first dose | 1 | 0 | 0 | 675 | 2,223 | 7 | 14,234 | 0.7 |
| ≥14 days after first dose until second dose | 1 | 1 | 0 | 408 | 1,506 | 5 | 12,112 | 0.8 |
| **Vaccinated with two doses** |  |  |  |  |  |  |  |  |
| 0-13 days after second dose | 0 | 0 | 0 | 162 | 483 | 11 | 4,751 | 0.0 |
| ≥14 days after second dose | 0 | 0 | 0 | 36 | 65 | 6 | 706 | 0.0 |

^*^We defined hospitalization related to a SARS-CoV-2 infection as a hospitalization that occurred within three days prior to or 14 days after an infection was initially confirmed. For attribution to person-days stratified by vaccination category, hospitalizations were assigned to the collection date for a confirmed infection.

**^†^**All deaths related to a SARS-CoV-2 infection were classified and confirmed by the California Correctional Health Care Services. For attribution to person-days stratified by vaccination category, deaths were assigned to the collection date for a confirmed infection.

#### Figure S7. Alternative model specifications to adjust for calendar time.

The admissions model with week-level fixed effects is shown in blue and the penalized spline with four degrees of freedom fit to calendar day is shown in grey.

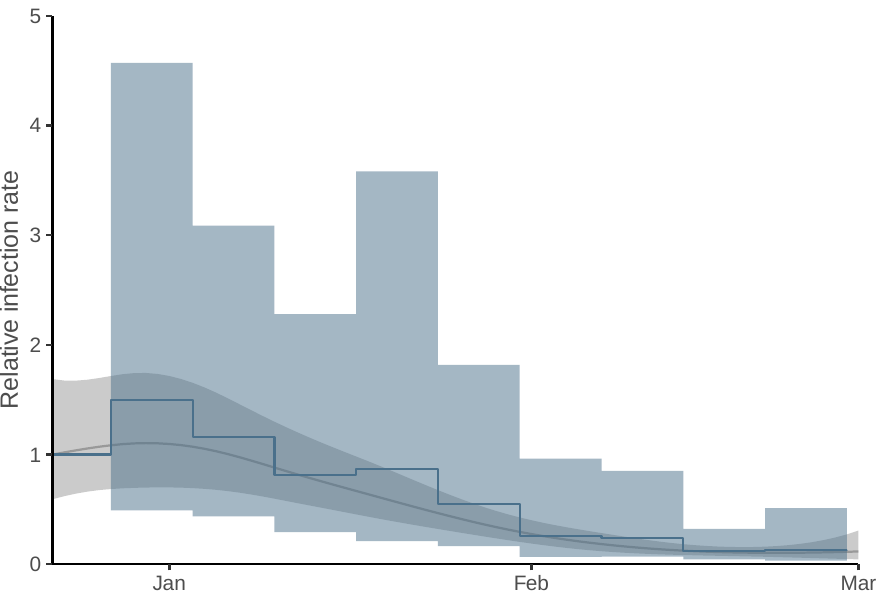

#### Table S6. Abbreviations for prisons in the California Department of Corrections and Rehabilitation system

| Prison | Acronym | County |
| --- | --- | --- |
| Avenal State Prison | ASP | Kings |
| California City Correctional Facility | CAC | Kern |
| California Correctional Center | CCC | Lassen |
| California Correctional Institution | CCI | Kern |
| California Health Care Facility | CHCF | San Joaquin |
| California Institution for Men | CIM | San Bernardino |
| California Institution for Women | CIW | Riverside |
| California Medical Facility | CMF | Solano |
| California Men's Colony | CMC | San Luis Obispo |
| California Rehabilitation Center | CRC | Riverside |
| California State Prison, Centinela | CEN | Imperial |
| California State Prison, Corcoran | COR | Kings |
| California State Prison, Los Angeles County | LAC | Los Angeles |
| California State Prison, Sacramento | SAC | Sacramento |
| California State Prison, Solano | SOL | Solano |
| California Substance Abuse Treatment Facility and State Prison, Corcoran | SATF | Kings |
| Calipatria State Prison | CAL | Imperial |
| Central California Women's Facility | CCWF | Madera |
| Chuckawalla Valley State Prison | CVSP | Riverside |
| Correctional Training Facility | CTF | Monterey |
| Deuel Vocational Institution | DVI | San Joaquin |
| Folsom State Prison | FSP | Sacramento |
| High Desert State Prison | HDSP | Lassen |
| Ironwood State Prison | ISP | Riverside |
| Kern Valley State Prison | KVSP | Kern |
| Mule Creek State Prison | MCSP | Amador |
| North Kern State Prison | NKSP | Kern |
| Pelican Bay State Prison | PBSP | Del Norte |
| Pleasant Valley State Prison | PVSP | Fresno |
| Richard J. Donovan Correctional Facility | RJD | San Diego |
| Salinas Valley State Prison | SVSP | Monterey |
| San Quentin State Prison | SQ | Marin |
| Sierra Conservation Center | SCC | Tuolumne |
| Valley State Prison | VSP | Madera |

### 5. STROBE statement

#### Table S7. STROBE checklist of items that should be included in reports of cohort studies

|  | Item No | Recommendation | Page No |
| --- | --- | --- | --- |
| **Title and abstract** | 1 | (*a*) Indicate the study’s design with a commonly used term in the title or the abstract | p.2 |
|  |  | (*b*) Provide in the abstract an informative and balanced summary of what was done and what was found | p.2 |
| Introduction | | | |
| Background/rationale | 2 | Explain the scientific background and rationale for the investigation being reported | p.4 |
| Objectives | 3 | State specific objectives, including any prespecified hypotheses | p.4 |
| Methods | | | |
| Study design | 4 | Present key elements of study design early in the paper | p.5 |
| Setting | 5 | Describe the setting, locations, and relevant dates, including periods of recruitment, exposure, follow-up, and data collection | pp.5-6 |
| Participants | 6 | (*a*) Give the eligibility criteria, and the sources and methods of selection of participants. Describe methods of follow-up | pp.5-6, & Supp. Append. s.I |
|  |  | (*b*) For matched studies, give matching criteria and number of exposed and unexposed | NA |
| Variables | 7 | Clearly define all outcomes, exposures, predictors, potential confounders, and effect modifiers. Give diagnostic criteria, if applicable | pp. 6-7 & Supp. Append ss.I, II |
| Data sources/ measurement | 8^*^ | For each variable of interest, give sources of data and details of methods of assessment (measurement). Describe comparability of assessment methods if there is more than one group | p.6 & Supp. Append s.II |
| Bias | 9 | Describe any efforts to address potential sources of bias | pp.7-8 & Supp. Append. S.II |
| Study size | 10 | Explain how the study size was arrived at | p.5-6, & Supp. Append. s.I |
| Quantitative variables | 11 | Explain how quantitative variables were handled in the analyses. If applicable, describe which groupings were chosen and why | Supp. Append. s.I |
| Statistical methods | 12 | (*a*) Describe all statistical methods, including those used to control for confounding | pp.6-8 & Supp. Append. s.II |
|  |  | (*b*) Describe any methods used to examine subgroups and interactions | pp.7-8 & Supp. Append. s.II |
|  |  | (*c*) Explain how missing data were addressed | Supp. Append. s.I |
|  |  | (*d*) If applicable, explain how loss to follow-up was addressed | NA |
|  |  | (*e*) Describe any sensitivity analyses | p.7-8 |
| Results | | |  |
| Participants | 13^*^ | (a) Report numbers of individuals at each stage of study—eg numbers potentially eligible, examined for eligibility, confirmed eligible, included in the study, completing follow-up, and analysed | p.8. Figure S1, Supp. Append. s.I |
|  |  | (b) Give reasons for non-participation at each stage | NA |
|  |  | (c) Consider use of a flow diagram | Figure S1 |
| Descriptive data | 14^*^ | (a) Give characteristics of study participants (eg demographic, clinical, social) and information on exposures and potential confounders | Table 1 |
|  |  | (b) Indicate number of participants with missing data for each variable of interest | Figure S1 |
|  |  | (c) Summarise follow-up time (eg, average and total amount) | p.8 |
| Outcome data | 15^*^ | Report numbers of outcome events or summary measures over time | p.9. Figure 1 & Table 2 |
| Main results | 16 | (*a*) Give unadjusted estimates and, if applicable, confounder-adjusted estimates and their precision (eg, 95% confidence interval). Make clear which confounders were adjusted for and why they were included | Table 2, Figure S6 & Supp. Append. s.2 |
|  |  | (*b*) Report category boundaries when continuous variables were categorized | NA |
|  |  | (*c*) If relevant, consider translating estimates of relative risk into absolute risk for a meaningful time period | NA |
| Other analyses | 17 | Report other analyses done—eg analyses of subgroups and interactions, and sensitivity analyses | pp. 10-11 & Table S3 & Supp. Append. s.II |
| Discussion | | | |
| Key results | 18 | Summarise key results with reference to study objectives | pp.11-13 |
| Limitations | 19 | Discuss limitations of the study, taking into account sources of potential bias or imprecision. Discuss both direction and magnitude of any potential bias | p.13-15 |
| Interpretation | 20 | Give a cautious overall interpretation of results considering objectives, limitations, multiplicity of analyses, results from similar studies, and other relevant evidence | pp.11-15 |
| Generalisability | 21 | Discuss the generalisability (external validity) of the study results | p.11-15 |
| Other information | | | |
| Funding | 22 | Give the source of funding and the role of the funders for the present study and, if applicable, for the original study on which the present article is based | p.18 |

^*^Give information separately for exposed and unexposed groups.

1. U.S. Census Bureau. 2010 census summary file 1— technical documentation. Washington, DC: U.S. Department of Commerce. SF1/10-4 (RV).
2. Witman A, Beadles C, Liu Y, et al. Comparison group selection in the presence of rolling entry for health services research: Rolling entry matching. Health Serv Res 2019;54(2):492–501.
3. Dagan N, Barda N, Kepten E, et al. BNT162b2 mRNA Covid-19 Vaccine in a Nationwide Mass Vaccination Setting. New England Journal of Medicine 2021;384(15):1412–23.
4. See e.g., 45 C.F.R. part 46, 21 C.F.R. part 56; 42 U.S.C. §241(d);  5 U.S.C. §552a; 44 U.S.C. §3501 et seq.
5. California Correctional Health Care Services. COVID-19 Testing Definitions and Strategies. COVID-19 and Seasonal Influenza: Interim Guidance for Health Care and Public Health Providers. Revised: February 19, 2021.
